# Selection of Human Single Domain Antibodies (sdAb) Against Thymidine Kinase 1 and Their Incorporation Into sdAb-Fc Antibody Constructs For Potential Use In Cancer Therapy

**DOI:** 10.1101/2021.05.04.21256554

**Authors:** Edwin J. Velazquez, Jordan D. Cress, Tyler B. Humpherys, Toni O. Mortimer, David M. Bellini, Jonathan R Skidmore, Kathryn R. Smith, Richard A. Robison, Scott K. Weber, Kim L. O’Neill

**Author notes:** Correspondence: Kim L. O’Neill, LSB 4007, Department of Microbiology and Molecular Biology, Brigham Young, University, Provo, UT, USA, 84602, Phone: 1+.

## Abstract

Thymidine Kinase 1 (TK1) is primarily known as a cancer biomarker with good prognostic capabilities for liquid and solid malignancies. However, recent studies targeting TK1 at protein and mRNA levels have shown that TK1 may be useful as a tumor target. In order to examine the use of TK1 as a tumor target, it is necessary to develop therapeutics specific for TK1. Single domain antibodies (sdAbs), represent an exciting approach for the development of immunotherapeutics due to their cost-effective production and higher tumor penetration than conventional antibodies. In this study, we isolated sdAb fragments specific to human TK1 from a human sdAb library. A total of 400 sdAbs were screened through 5 rounds of selection by monoclonal phage ELISA. The most sensitive sdAb fragments were selected as candidates for preclinical testing. The sdAb fragments showed specificity for human TK1 in phage ELISA, Western blot analysis and had a limit of detection of 3.9 ng/ml for 4-H-TK1_A1 and 1.9 ng/ml for 4-H-TK1_D1. The antibody fragments were successfully expressed and used for detection of membrane associated TK1 (mTK1) through flow cytometry on cancer cells [lung (∼95%), colon (∼87%), breast (∼53%)] and healthy human mono nuclear cells (MNC). The most sensitive antibody fragments, 4-H-TK1_A1 and 4-H-TK1_D1 were fused to an engineered IgG1 Fc fragment. When added to cancer cells expressing mTK1 co-cultured with human MNC, the anti-TK1-sdAb-IgG1_A1 and D1 were able to elicit a significant antibody-dependent cell-mediated cytotoxicity (ADCC) response by human MNCs against lung cancer cells compared to isotype controls (*P*<0.0267 and *P*<0.0265, respectively). To our knowledge this is the first time that the isolation and evaluation of human anti TK1 single domain antibodies using phage display technology has been reported. The antibody fragments isolated here may represent a valuable resource for the detection and the targeting of TK1 in tumor cells.

## Introduction

Efficient DNA repair and synthesis requires a balanced supply of nucleotides and coordination of the metabolic pathways utilized for their production [1]. In order to sustain proliferation, malignant cells have significant alterations in the activity levels of several of their nucleotide synthesis enzymes [2-4]. These alterations, particularly in the pyrimidine salvage pathway, can lead to an imbalance in the cell’s nucleotide pools which could lead to error prone DNA replication and genome instability, a hallmark of cancer [5-7]. Thymidine Kinase 1 (TK1) is a pyrimidine salvage pathway enzyme that catalyzes the phosphorylation of thymidine to thymidine monophosphate [8]. In healthy cells, TK1 is only elevated during the S phase, but low or absent during other cell cycle stages [9-10]. However, in malignant cells TK1 expression levels are upregulated and the enzyme seems to lose its normal cell cycle regulation control elements [11]. As cancer progresses, TK1 activity levels increase in tissues and in serum proportionally to tumor size and stage of disease [12-13]. During the last 3 decades scientific evidence has shown that TK1 levels in the serum of cancer patients can be used as a biomarker for early cancer detection [14-16]. As TK1 levels in serum are correlated with tumor progression, patient response and cancer recurrence, TK1 has also been proposed as a suitable tumor biomarker for the continued monitoring of patients [17-20].

Although the usefulness of TK1 as a tumor biomarker has been the main focus of many studies, in recent years the interest in using TK1 as a tumor target for multiple cancers has gradually increased [21]. In a study conducted by Malvi et al., the silencing of TK1 in lung adenocarcinoma (LUAD) cell lines inhibited the cell growth, migration and invasion capacities of LUAD cells both in vitro and in vivo [22]. Similarly, another study showed that targeting of TK1 through genetic knockdown significantly reduced cell proliferation of pancreatic ductal adenocarcinoma (PDAC) cells [23]. In addition, it has been reported, that some forms of TK1 seem to be able to associate to the cell membrane of several cancer cell types, including leukemia, breast, lung and colon tumor cells possibly through protein-protein interaction or transitory membrane localization through exosomes [24-26]. Moreover, early experimental data has shown that membrane associated TK1 (mTK1) in lung, colon and breast cancer cells could be targeted using monoclonal antibodies. However, the study was limited in some extent because the antibodies used were produced in mice and humanized antibodies are still required to better evaluate potential therapeutic use in humans [27]. This evidence together suggests that the targeting of TK1 both inside the cell and its mTK1 form could be a possible approach for the development of novel cancer therapies. Therefore, the generation of therapeutics specific for TK1 could enable us to explore the potential of TK1 as a tumor target.

Monoclonal antibodies (mAbs) are suitable candidates for the development of cancer therapies due to their high specificity and affinity for their molecule targets [28]. The majority of current therapeutic antibodies have been produced with hybridoma technology or in transgenic mice, these approaches require the use of special animals, time consuming protocols and humanization or reformatting through complicated techniques such as CDR engraftment before any therapeutic use is possible [29, 30].

Recently, phage display technology has been incorporated in the production pipeline of therapeutic antibodies by many pharmaceutical companies [31, 32]. This technology offers the possibility to explore vast human antibody libraries in a relatively short period of time compared to hybridoma technology and does not require animals and can isolate antibodies against low immunogenic antigens [33]. In addition, the antibody fragments can be isolated in convenient formats that facilitate further modifications for therapeutic applications [34, 35]. The use of phage display technology to obtain single domain antibody fragments (sdAbs), also called nanobodies, against TK1 is a convenient and appealing approach to isolate and develop biopharmaceuticals specific for the targeting of TK1. In this study we isolated human antibody fragments against the tumor proliferation biomarker TK1 from a sdAb library. The antibody fragments were evaluated for their capacity to bind and detect TK1 in monoclonal phage ELISA, Western blot and flow cytometry. The antibody fragments were then incorporated into engineered IgG1 constructs and tested for their capacity to target TK1 in malignant cells and elicit an ADCC response from human MNCS against cancer cells. We hypothesize that engineered single domain antibodies (sdAb) specific for TK1 can efficiently target TK1 high-level expressing tumor cells. Thus, the use of human sdAb molecules targeting TK1 may enable us to better explore the potential of TK1 as a tumor target in proliferating malignant cells.

## Materials and methods

### Isolation of anti-TK1 sdAb fragments through phage display

A phage display library of human single domain antibodies developed by Dr. Daniel Christ at the MRC laboratory of molecular biology was used to select high affinity anti-TK1 sdAbs (Source Bioscience, Cambridge, U.K) as previously described [36]. The full repertoire of sdAbs was contained within a single human VH framework (V3-23/D47) fused to the gene III protein of the M13 filamentous phage. The sdAb library was constructed into the pR2 (MYC VSV-G tag) plasmid and had a diversity of 3×10^9^ fragments. Each sdAb fragment contained three diversified complementary regions (CDR1, CDR2 and CDR3). Ubiquitin and Galactosidase sub libraries were also included as positive controls and run previously according to the manufacturer instructions to test the viability of the components of the library and verify the selection process and testing of the sdAbs were performed properly.

To initially amplify the full repertoire of sdAbs, one aliquot of the library was grown in 500 mL of 2xTY media supplemented with 4% glucose and 100ug/ml of ampicillin until the culture reached an OD_600_ of 0.5. An amount of 1×10^12^ KM13 helper phages were added per each 250 ml of culture, and the culture was incubated for 1 hr. at 37 °C without agitation. After infection, the media was replaced with 2xTY media containing 0.1 % glucose, 100 μg/ml of ampicillin and 75 μg/ml of kanamycin and the library was grown for 24 hr. at 25 °C on a shaker at 250 rpm. The phages displaying the sdAb fragments were then purified using polyethylene glycol 6000 (PEG) solution (Millipore SIGMA, St Louis, MO, USA). The purified phages were then quantified infecting TG1 bacteria with serial dilutions of the phage and plating the infected TG1 on TYE amp plates. Full-length human recombinant TK1 (>80% pure) produced in *E coli* (Genscript, Piscataway, NJ) was diluted in phosphate buffered saline (PBS) buffer at a 0.05 mg/ml concentration. TK1 was then immobilized on Maxisorp plates (ThermoFisher scientific, Waltham, MA), by coating four wells with 100 ul of TK1 solution and incubating them at 4 °C overnight. After coating the plates overnight, the plates were blocked with 5% milk PBS buffer (MPBS) for 45 min at room temperature on a shaker and 15 minutes at 37 □C. The plates were then washed 3 times with PBS. Approximately 5×10^10^ phages displaying the full repertoire of sdAbs, were applied to each well that was coated with TK1. Phages were allowed to bind for 1 hr at room temperature with moderate shaking. After incubation, the wells were washed 15-20 times with PBS-T buffer (0.1% Tween) and 2 times with PBS buffer. The phage-sdAbs that remained attached were then eluted adding 100 μL of a 0.1 mg/mL trypsin-TBSC buffer solution (Millipore SIGMA, St Louis, MO, USA) and incubated 1 hr at room temperature with moderate shaking. The eluted phages were then recovered and used to infect a 30 mL TG1 bacteria culture at 0.5 OD600. The infected culture was then incubated for 1 hr at 37 □C without shaking. After infection, the TG1 bacteria was harvested and resuspended in 1 mL of 2xTY media. The cells were then plated on TYE, 4% glucose plates with ampicillin (100μg/ml). The next day, colonies were scraped off and grown in 500 mL of 2xTY until the cultures reached at 0.5 OD_600_. The cultures were then infected with 1×10^12^ KM13 phages and grown at 25 □C for 24 hr on a shaker at 250 rpm. After incubating for 24 hr, the phage-antibody sub library was purified using PEG 6000 and the phage titter was determined for both the eluted phages and the purified phages. This process of selection was repeated 5 times. To eliminate antibodies that could possibly bind to 6xHis-tag in TK1, the last two rounds of selection were done using TK1 produced in HEK 293expi cells without 6xHis-tag (Origene, Rockville, Maryland, USA).

### Polyclonal and Monoclonal Phage ELISA

To monitor the increase in the overall number of TK1 binders between rounds of selection the isolated phages from each round of selection were screened through polyclonal phage ELISA. For polyclonal phage ELISA 96-well Costar plates (Corning, NY, USA) were coated with 100 μl of serial dilutions of TK1 (0.05 mg/ml-0.0004 mg/ml) overnight at 4°C with gentle agitation. After coating the wells, plates were washed 3 times with PBS buffer and blocked with 240 μl of MPBS for 45 minutes at room temperature and 15 minutes at 37°C. Wells were washed 3 times with PBS and purified phages diluted in MPBS (1:1 ratio) were added into each well. The plates were incubated for 1 hr at room temperature with gentle agitation. After incubating the plates were washed with PBS-T five times and 100 μl of Horse Radish Peroxidase (HRP) conjugated anti-M13 antibody solution (1:2,000 dilution in MPBS) were added into each well. Following addition of the HRP conjugated antibody, 100 μl of Tetramethyl Benzidine (TMB) substrate (ThermoFisher scientific, Waltham, MA, USA) was added into each well and color was allowed to develop 10-30 min. The reaction was stopped with 50μl of 1M sulfuric acid, and the absorbance values were measured using a Synergy HT Microplate Reader (Bio-Tek Winooski, VT) at 450 nm and 650 nm.

Eighty individual clones were tested using monoclonal phage ELISA every round of selection (biopans). After each biopan and before scraping off the colonies, eighty clones were picked and grown overnight at 37°C in 200 μl of 2XTY media supplemented with 4% glucose and ampicillin (100 μl/ml). Culture dilutions (1:100) from the overnight cultures were made by diluting 5 μl of the overnight culture in 200 μl of 2XTY 4% glucose and ampicillin and grown at 37°C for three hr. After reaching a 0.5 OD_600_, 50 μl of 2XTY containing 4 x 10^8^ KM13 helper phages were added into each well to produce phage-sdAb fragments. The infected cultures were incubated at 37°C for one hr without shaking and the media was changed with 200 μl of 2XTY with 0.1% glucose, ampicillin (100 μg/ml), and kanamycin (75 μg/ml). Cultures were grown at 25°C for 24 hr at 250 rpm. After 24 hr the plates were centrifuged at 3200 xg for 10 minutes and the supernatants were recovered from each well and mixed with MPBS in 1:1 ratio. To test each clone for its capacity to bind TK1 96 well plates were coated with 100 μl of a 0.01 mg/ml of TK1 per well as previously described. The same protocol described for polyclonal phage ELISA was used to carry out the monoclonal phage ELISA.

### Detection of phage-sdAbs by dot blot

Dot blot was used to confirm the expression of sdAb fragments in TG1 supernatant. After recovering supernatants containing phage-sdAb fragments, 2-3 μl of the phages were immobilized on a (nitrocellulose membrane BIO-RAD, Hercules, CA). The membrane was then blocked with 5% MPBS for 1 hr at room temperature on a shaker. The membrane was then incubated with a 1:20,000 anti-VSV-G-HRP antibody solution at 4 °C overnight with moderate shaking. The next day, the membrane was washed 3 times with PBS-T 5 min each wash. After the membranes were washed 2 mL of enhanced chemiluminescence substrate (Advansta Corporation, San Jose, CA) was added to the membrane until the membrane was completely covered. The membranes were incubated for 2 minutes and the excess of reagent was poured off. Membranes were covered in plastic wrap and light sensitive films were placed on the membranes at different exposure times and revealed using an imaging developer.

### Sensitivity of TK1 specific antibody fragments

Sensitivity of TK1 sdAbs was determined using dose-response curves and monoclonal phage ELISA. Briefly, Costar 96-well plates (Corning) were coated with serial dilutions of TK1 (ranging from 23,600 ng/ml-23 ng/ml for E-TK1 and 500 ng/ml-1.9 ng/ml for H-TK1) overnight at 4°C overnight. The next day, plates were blocked with 2400 μl of MPBS/well for 45 minutes at room temperature and 15 minutes at 37°C. After blocking, the wells were washed three times with PBS and 100 μl of a 1:1 dilution of phage supernatant in MPBS was added into each well. Subsequent steps were carried out as previously described to develop the ELISA. The curves were then analyzed using a four-point parameter logistic curve and the limit of detection of each sdAb fragment was determined. The sensitivity of the clones was then compared. In the case of soluble fragments that were expressed without fusion to the PIII protein of the phage, the same ELISA protocol was used except detection antibodies changed to anti-His-HRP (Biolegend, San Diego, CA) and anti-VSV-G-HRP (Bethyl, Montgomery, TX).

### Validation with a TK1 siRNA and non-specific binding controls

In order to confirm specific binding to TK1, individual clones were screened against cell lysate from a siRNA TK1 knockout and compared to wild type cell lysate. TK1 knock down cell lysate was prepared as previously described [25]. TK1 produced in bacterial, yeast, and mammalian expression systems were used as positive controls and uncoated wells were used as negative controls. The protocol for a monoclonal phage ELISA was performed as described above.

### Sequencing Analysis of sdAb fragments

Plasmid was isolated from the clones that showed the highest affinity for TK1 using the PureYield Plasmid Miniprep system (Promega). Samples were prepared for sequencing using the primer (5’ CCCTCATAGTTAGCGTAACGA 3’) and the universal M13 reverse primer (5’ CAGGAAACAGCTATGAC 3’). Sequencing data was analyzed using Genious prime software [37].

### Anti-TK1-sdAbs protein modeling and docking analysis of sdAb-TK1complexes

The structures of the anti-TK1-sdAb fragments were analyzed using the GalaxyWEB TMB web server. The most stable structures of each anti-TK1 sdAb fragment were analyzed using the Visual Molecular Dynamics (VMD) software developed by the computer science, and biophysics at the university of Illinois [38]. The CDR regions were mapped by analyzing the deduced amino acid sequences of each anti-TK1-sdAb fragment in the IgG Blast tool from NCBI. The sequences of the anti-TK1-sdAb fragments were aligned using Genious prime software. In silico analysis of the interaction between the anti-TK1-sdAb fragments and the crystal structure of TK1 was performed using the high ambiguity driven protein-protein docking (HADDOCK) web server [39]. Visualization of the anti-TK1-sdAb-TK1 complexes was also performed with VDM software.

### PCR amplification and cloning into pET-scFv-T

Sequences of the sdAb fragments were amplified from the phagemid plasmids corresponding to the isolated positive clones in phage ELISA. The sequences were amplified using primers containing the NcoI restriction site (5’ GAA<u>CATATG</u>ATGAAAAAATTATTA 3’) and the NotI restriction site (5’ GAA<u>GGATCC</u>TGCGGCCCCCTTTC 3’). PCR products were run in a 1% agarose gel and desired sequences were extracted using the Zymoclean DNA Gel Recovery Kit (Zymo research, Irvine, CA, USA). Following gel recovery, sdAb sequences were digested using NcoI and NotI restriction enzymes (New England Biolabs, Ipswich, MA, USA). Digested sequences were then ligated into the pET-scFv-T backbone (Addgene, Watertown, MA). Ligation was carried out using the Quick Ligation kit (New England Biolabs, MA). The ligated plasmids were then cloned into Dhα5 competent cells. Colonies from transformation product were grown for 16-20 hr and plasmid were isolated and analyzed by restriction enzyme analysis to verify the presence of the insert. Positive clones were then sequenced.

### Expression of antibody fragments in Rosseta 2(DE3) pLysS E. coli cells and His-tag purification

The pET-TK1-sdAb-6xHis constructs were cloned into Rosseta blue(DE3) pLysS *E. coli* cells (Millipore SIGMA, St Louis, MO). Individual colonies were selected and grown in a culture overnight at 37°C. The overnight culture was then scaled up and grown until the OD reached 0.6. Expression of the sdAb fragments was induced by addition of 0.4 mM Isopropyl β-d-1-thiogalactopyranoside (IPTG). After IPTG induction, the cultures were grown for 24 hr at 28°C. The Rosseta blue(DE3) pLysS *E. coli* cells were pelleted and the supernatant was saved for later purification. The cells were subjected to osmotic shock by resuspending cells in TES buffer (20 mM Tris-HCl pH 7.6, 5 mM EDTA, and 20% sucrose). After a one hr incubation on ice, the sample was centrifuged at 14,000 xg for 20 minutes. The pellets were resuspended in ddH_2_O and incubated on ice for 30 minutes. Centrifugation was repeated and the supernatant was preserved. The cell pellet was lysated using 10X bug buster reagent (Millipore SIGMA, St Louis, MO) diluted in 20 mM Tris–HCl (pH 7.8) buffer containing 15 mM NaCl, 5 mM MgCl2, DNase (25 U/ml) and protease inhibitors. Cells were lysated for 20 min on shaker with moderate agitation. After incubating in cell lysis, the lysated cells were spun down 16,000 g for 20 min and the supernantant was recovered. Supernatants were mixed with equilibrated Ni-NTA agarose beads (Qiagen, Hilden, Germany) for 3 hr at 4 °C. After incubating the NI-NTA beads were washed twice with cell lysis buffer and placed into 5 ml polyproylene columns. The Ni-NTA beads were then washed with 50 ml of wash buffer and then the His-tagged proteins were eluted with elution buffer in 0.3 ml fractions. The fractions were analyzed by SDS-PAGE and Western blot to detect the purified anti-TK1 sdAb fragments and estimate their purity.

### Western blot with purified anti TK1-sdAb fragments

The purified fragments were tested for their capacity to bind to purified TK1 and TK1 in cell lysate through Western blot. Recombinant human TK1 produced in bacteria and Expi293F cells were used along with cell lysate form A549 lung cancer cell lysates, including a siRNA TK1 knockdown cell lysate. Briefly, 0.5 μg of TK1 or 20 μg of cell lysate were mixed with 6x Laemmli buffer (Millipore SIGMA, St Louis, MO). The protein samples were then heated at 100 °C for 5 min and loaded into a 12 % SDS-PAGE electrophoresis. The proteins from the gel were then transferred to nitrocellulose membranes (Bio-Rad, Hercules, CA, USA). After blocking with 5 % milk in MPBS buffer for 1 hr at room temperature the blocking solution was poured out and anti-TK1 sdAb fragment solution (1 ug/ml-2 ug/ml) was added. Membranes were incubated at 4 °C overnight. After overnight incubation membranes were washed 3 times with PBS-T buffer. The bound proteins were detected using a 1:20,000 solution of an anti-VSV-G-HRP and anti-His-HRP antibody (Bethyl, Montgomery, TX, USA). The proteins were then detected through the peroxidase reaction using enhanced chemiluminescence (ECL) (Advansta Corporation, San Jose, CA). Films were exposed for different amounts of time depending on the antibody being tested, times ranged from 30 seconds to 5 min. The films were scanned, and the images were analyzed using the software ImageJ from NIH.

### Cell lines and Isolation of human MNCs

The NCI-H460 (ATCCHTB-177™), A549 (ATCC^®^ CCL-185), HCC1806 (ATCC^®^ CRL-2335™) and HT-29 (ATCC^®^ HTB-38™) cell lines were obtained from the American Type Culture Collection (ATCC, Manassas, VA, USA) and maintained according to ATCC recommendations. A549, NCI-H460, HCC1806 and HT-29 were cultured in RPMI-1640 media (ThermoFisher scientific, Waltham, MA) supplemented with 10% fetal bovine serum (FBS) and 2mM L-Glutamine. All cell lines were grown in an incubator at 37 °C and 5% CO_2_. All cell lines used were tested for TK1 surface expression with flow cytometry with a commercial antibody Abcam91651 (Abcam, Cambridge, UK) to confirm the presence of TK1 on the cell membrane. MNC were isolated with lymphocyte separation media (Corning, NY) following manufacturer instructions, red blood cells were depleted with red blood cell lysis buffer (Biolegend, San Diego, CA) and the MNC were resuspended in LGM-3 (Lonza, Basel, Switzerland). Blood withdrawal was done under the institutional review board approval.

### Flow cytometry

The purified phage-sdAb fragments, anti-TK1 sdAb fragments, and anti-TK1-sdAb-IgG1 fusions were all tested for their capacity to detect mTK1 in the cancer cell lines NCI-H460, HCC1806, HT-29, and normal MNCs. For this analysis 1×10^6^ cells per sample were analyzed. Cells were washed twice with PBS and resuspended in 200 μl of cell staining buffer. After resuspending, the cells were stained using PEG purified phage-TK1-sdAb fragments (100 μl), purified sdAb fragments (5-10 ug), or purified sdAb-IgG1 antibodies for 40 min. The cells were then washed 3 times with 200 μl of cell staining buffer and stained for 30 min with anti-his-APC or anti-VSV-G-FITC antibodies in the case of purified phage-sdAb fragments, anti-M13-FITC secondary antibody. To detect binding of sdAb-IgG1 antibody fusions anti-Human IgG-FITC antibody (Abcam, Cambridge, UK) was used. After incubation with secondary antibody, the cells were washed 3 times with 200 μl of cell staining buffer. Before analysis samples were stained with 10 μg/ml PI solution and the samples were analyzed in a Cytoflex flow cytometer machine (Beckman Coulter, Brea, CA). The FCS files were analyzed using the FlowJo sowtware (FlowJo, Inc., Ashland, OR).

### Incorporation of TK1-sdAb fragments into pFUSE-IgG1 construct and antibody expression in CHO cells

The DNA sequences of the top two anti-TK1 sdAbs were cloned between the NcoI and EcoRI restriction sites of the pFUSE-hIgG1e5-Fc2 vector (InvivoGen, San Diego, CA). This vector is designed for the production of human recombinant antibodies in mammalian cells and contains a human IgG1 heavy chain mutated at the S239D/A330L/I332E sites which confers an increased binding to FcγIIIa receptors in macrophages (MO) and natural killer cells (NK). Thus, the recombinant antibodies fused to this engineered IgG1, can elicit an enhanced antibody-dependent cell-mediated cytotoxicity (ADCC). The primers used for the amplification of the sdAb fragments for this construct were primer Fw-5’ GAAGAATTCGATGGCCGAGGTGCAG 3’ and primer Rv-5’ GGCCCATGG CGCTCGAGACGGTGAC 3’. The two TK1-scFv-hIgG1 DNA constructs were introduced into CHO.K1 cells (ATCC, Manassas, VA, USA) by lipofection using the lipofectamine LTX reagent (ThermoFisher scientific, Waltham, MA). After 48hr the media was changed and Zeocin selection antibiotic (InvivoGen, San Diego, CA, USA) was added at a concentration of 100 μg/ml. After 10 days in selection the cells were expanded. The media was then changed with ProCHO™ AT (Lonza, Basilea, Switzerland) and the recombinant antibodies were allowed to be produced for 48-96 hr or until cell viability was about 50 %. The media was collected and cleared from cells by centrifugation. The anti-TK1-scFv-hIgG1 antibodies where then purified from cleared media using protein A purification columns (ThermoFisher scientific, Waltham, MA). Characterization of the purified recombinant antibodies was carried out with Western blot and flow cytometry as described above.

### In vitro testing of anti-TK1-sdAb-IgG1 Antibodies through ADCC

The capacity of the TK1-sdAb-IgG1 antibodies to target mTK1 on cancer cells and elicit an ADCC response was evaluated *in vitro*. For this experiments NCI-H460 cells, which expressed high levels of mTK1, were engineered to express cytosolic GFP. The cells were then co-cultured with human MNC and anti TK1-sdAb-IgG1 antibodies were added in various concentrations. Cell death was then measured using a real time cell imaging system. The experiments were conducted as follows. One day before treating the cells with antibodies and controls 5000 NCI-H460 GFP+ cells were seeded per well in a 96-well tissue culture plate (MIDSCI, St. Louis, MO), placed inside an ImageXpress^®^ Pico system. The GFP+ cells were counted every hr for the next 8-12 hr. After initial growth human MNCs were added at two different ratios, 5:1 and 10:1. The cells were co-cultured using LGM-3 media to sustain MNC and antibodies were added at various concentrations (20, 10, 5, 2.5 ug per well). And optimized 5:1 effector:target ratio and a concentration of 10 μg of antibody/ml were used for experiments. The cells were monitored for 72-96 hr under environmental controlled conditions. The number of cells from each treatment including controls were analyzed and compared through time. Each experiment was run twice and each well receiving a treatment was run in duplicate.

### Statistical analysis

Statistical analyses were performed using the GraphPad Prism software (GraphPad, San Diego, CA). ELISA data from dilution curves was log-transformed and analyzed with a 4-parameter non-linear regression analysis with a 95% confidence interval (CI). To compare the different treatments of the ADCC experiments, the data was normalized in reference to the moment MNC and antibodies were added and analyzed using a two-way ANOVA with repeated measure analysis. Analysis of multiple comparisons was performed comparing the mean of each treatment with every other treatment mean in each time point and over the total course of time. One-way ANOVA were performed to compare normalized GFP+ cell counts at specific time points.

## Results

### Antigen validation

Each of the antigens used for the selection of the sdAbs were validated before the biopanning process. Recombinant human TK1 was produced in *E. coli* (E-TK1) and was used during early rounds of selection at high concentrations while TK1 produced in human Expi293F cells (H-TK1) was used for the last rounds of selection at lower concentrations. This is because H-TK1 that produced in human cells was properly folded. Both antigens were validated through Western blot using the KO validated anti-TK1 antibody ab91651(Abcam, Cambridge, UK). Both E-TK1 and H-TK1 purified fractions were positive for TK1 and showed bands respective to the monomer and dimer of TK1. The pureness of the antigens showed to be higher than 80% according to SDS page and Coomassie blue staining (Fig. 1A, right).

**Fig. 1.**
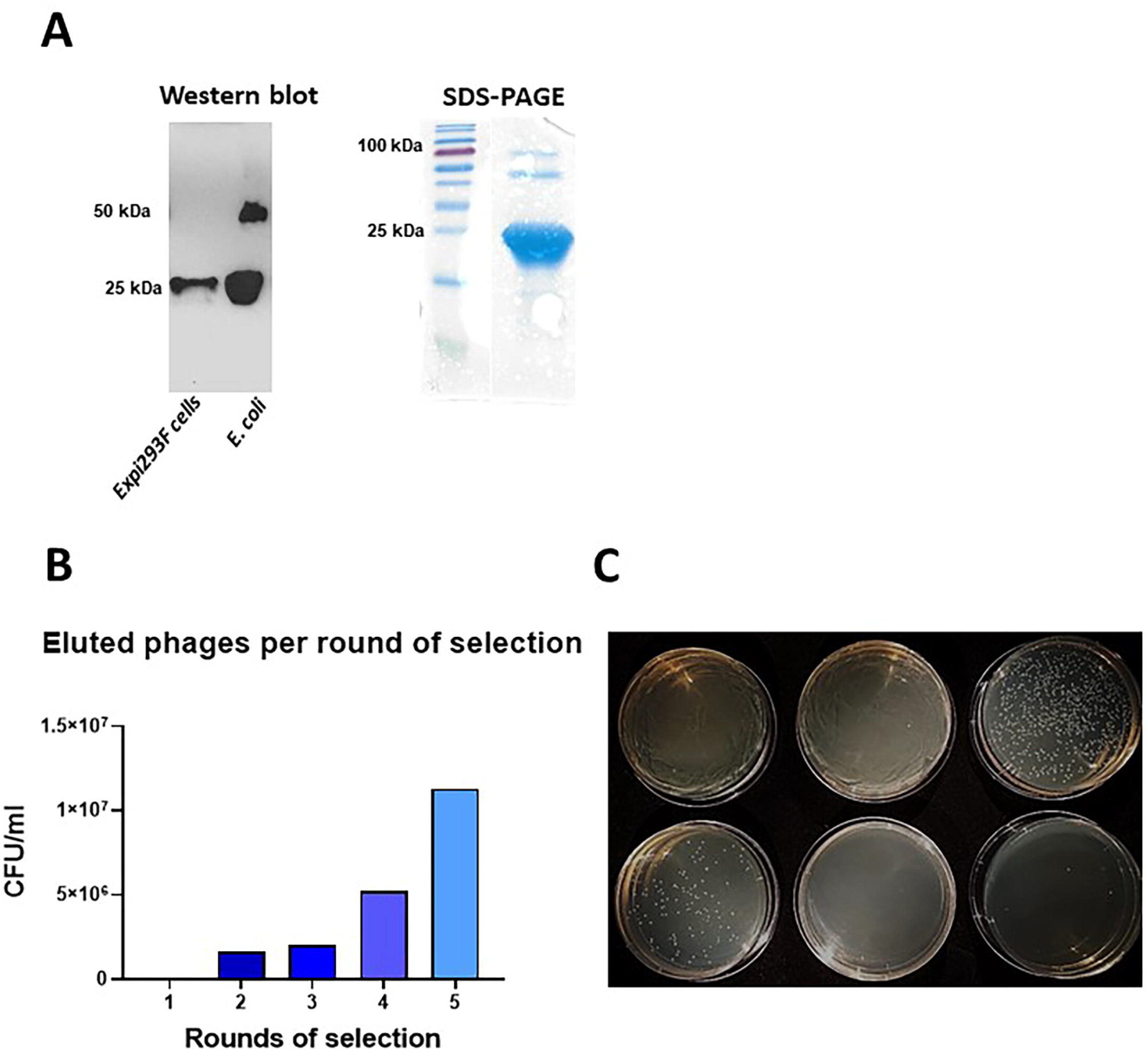
A) Antigen validation, the pureness and integrity of human recombinant TK1 produced in *E. coli* cells and Expi293F cells was assessed in SDS-PAGE and validated through Western blot using the anti-TK1 antibody ab91651. B) Enrichment of TK1 binders through 5 rounds of selection. The number of binders was estimated based on titrations of eluted phages used to infect TG1 bacteria. C) A representative image of a viral titration to determine the number of eluted phages. TG1 bacteria was infected with serial dilutions of the eluted phages after each biopans. The infected TG1 was then plated on TYE amp plates.

### Isolation and validation of anti-TK1 sdAb fragments

The isolation of anti-TK1-ssdAbs was monitored after each biopan by determining the phage titer from the eluted phages. As expected, the number of eluted phages per ml increased exponentially through the 5 rounds of selection (Table 1, Fig. 1B). This trend could be observed by infecting TG1 bacteria with the eluted phages, plating the bacteria in selective agar plates and then determining the number of CFU/ml (Fig. 1C). After 3 consecutive rounds of selection with recombinant human TK1 produced in *E. coli* the enrichment factor of TK1 binders went from 1 to 48.9. (Table 1). Two more rounds of selection were performed using recombinant H-TK1 to eliminate non-specific sdAb fragments that could possibly bind to the His-tag present in E-TK1 and also to obtain fragments that could bind to properly folded human TK1. We observed that the enrichment factor increased in a 3-fold after 1 round of selection with H-TK1 and then doubled in a subsequent round of selection using H-TK1.

**Table I.** Enrichment of anti-TK1-sdAb phages through 5 rounds of selection.

| Round of selection | Input phages | eluted phages | Ration (eluted/input) | Enrichment factor |
| --- | --- | --- | --- | --- |
| 1 | 5.00E+12 | 4.12E+04 | 8.24E-09 | 1.00 |
| 2 | 5.00E+12 | 1.60E+06 | 3.20E-07 | 38.83 |
| 3 | 5.00E+12 | 2.01E+06 | 4.03E-07 | 48.91 |
| 4 | 5.00E+12 | 5.20E+06 | 1.04E-06 | 126.21 |
| 5 | 5.00E+12 | 1.13E+07 | 2.27E-06 | 275.49 |

After each biopan 80 individual clones were screened through monoclonal phage ELISA. The purified phages after each round of selection were also monitored through polyclonal phage ELISA. As expected, an exponential increase in the number of positive clones after each biopan was observed. About 50 % of the clones produced a positive signal by the 4^th^ biopan and about 90% of the clones were positive in the 5^th^ biopan (Fig. 2A). A similar trend was observed in polyclonal phage ELISA where the signal produced by the total purified phages after each round of selection also significantly increased after the first biopan and kept increasing almost in a 2-fold between each biopan from the 2^nd^ to the 4^th^ biopans (Fig. 2B). Although the number of positive clones in monoclonal phage ELISA showed a significant increase from the 4 to the 5^th^ biopan, no significant increase in the overall signal was produced in polyclonal phage ELISA. This was consistent with what was observed in the monoclonal phage ELISAs where we see significant increases in the number of positive clones from the 1^st^ through the 4^th^ biopans.

**Fig. 2.**
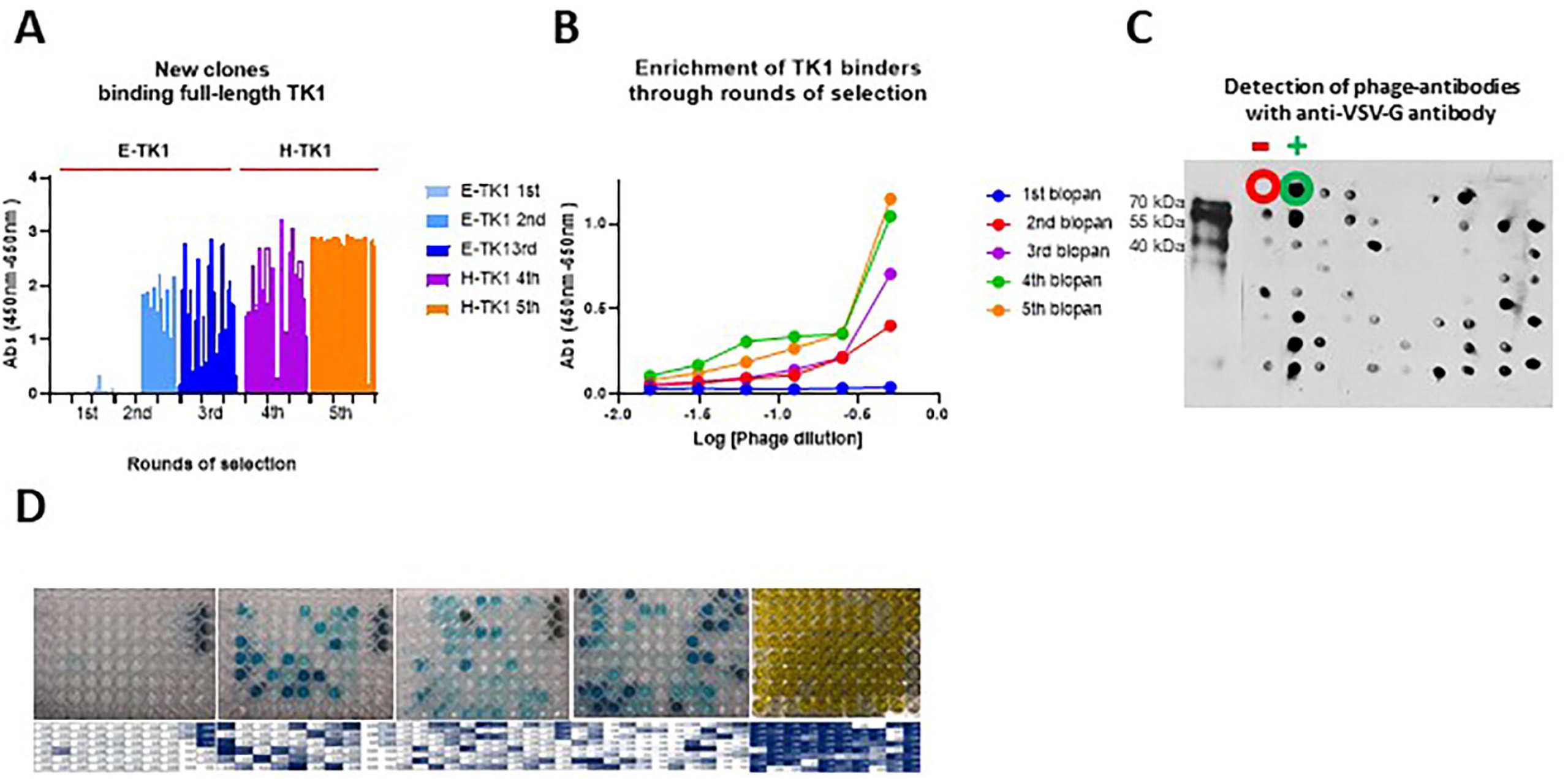
Selection and expression of anti-TK1-sdAb phages. A) Analysis of 80 clones through monoclonal phage ELISA was done after each round of selection. B) Polyclonal phage ELISA using the total purified phages per round of selection. In both A and B and increase in the overall signal and number of positive clones increases after each round of selection. C) Detection of anti-TK1-sdAb phages through dot blot using an anti-VSV-G-HRP antibody. Also, verification of packaging of the library from PEG purified phages after initial amplification of the sdAb library. D) Representative image of monoclonal phage ELISAs through the rounds of selection.

In addition to monoclonal and polyclonal phage ELISA, we screened purified phages and individual supernatants containing phages with Western blot and dot blot. Since each antibody fragment that is displayed in the M13 phages has a Myc and VSV-G tags we detected the production of phage-antibodies using anti-VSV-G-HRP conjugated antibody. Western blot analysis of the purified phages revealed the presence of bands corresponding to phage-sdAb fusions. Dot blot analysis of individual clones showed the successful production of individual phage-sdAbs (Fig. 2C).

After the initial screens in monoclonal phage ELISA, 26 clones were chosen for their ability to produce high signals. These included clones from the 2^nd^, 3^rd^ and 4^th^ rounds of selection. Clones from the 5^th^ round were excluded due to a decrease in the diversity of sequences product of amplification of some specific phage-sdAbs with high affinity. The clones were re-tested to confirm their capacity to bind TK1 and reproduce a positive signal. From the 26 selected clones, 14 clones were able to reproduce a positive signal after the initial screen (Fig. 3).

**Fig. 3.**
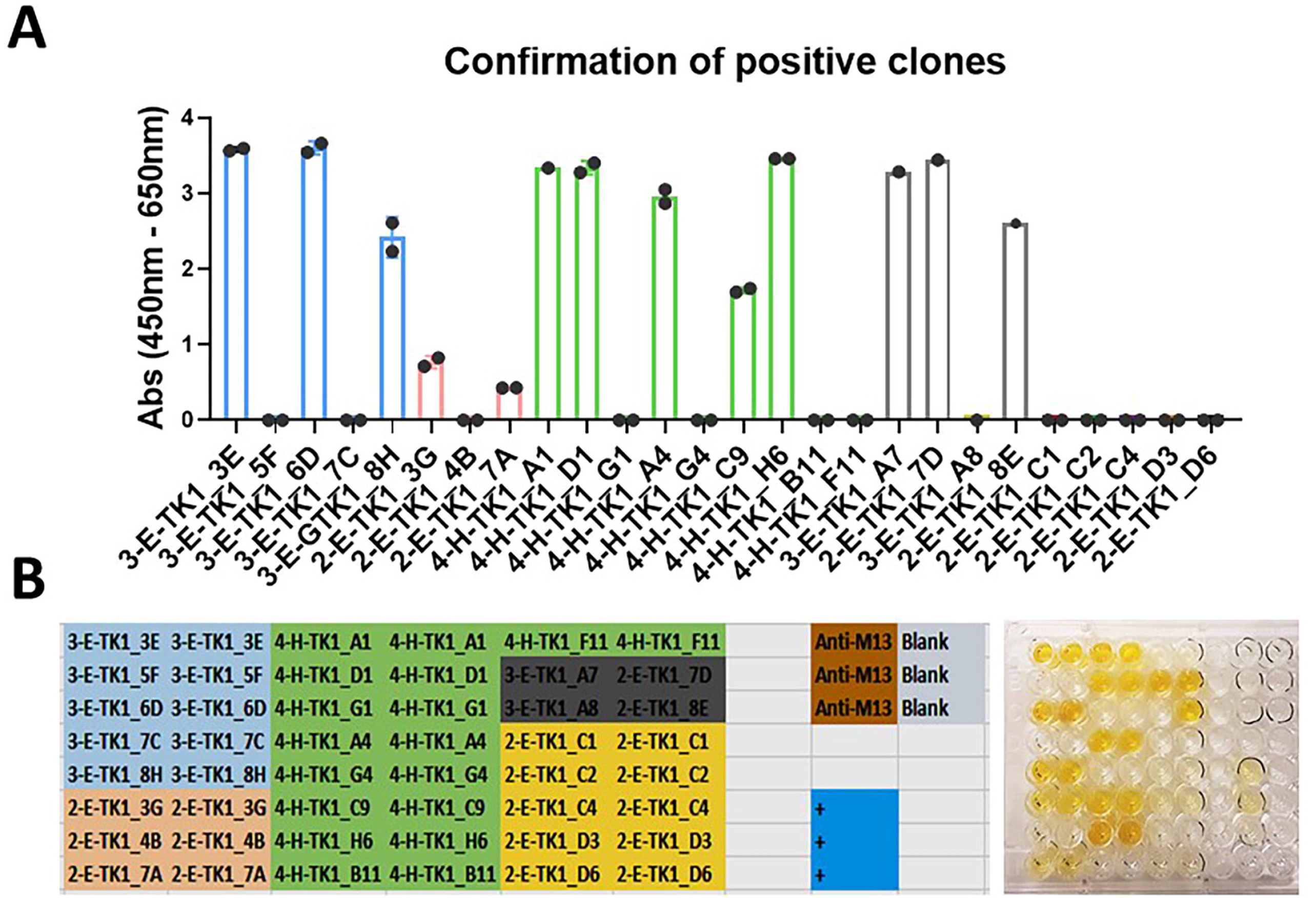
Confirmation of positive clones. To verify that the positive clones found during the initial screening, their capacity to reproduce a positive signal was tested. The strongest positive clones were selected through the different rounds of selection. Bacteria corresponding to each of these clones was streaked for second time on TYE amp plates. New cultures were grown from a single colony and infected with KM13 helper phage to induce the production of their respective anti-TK1 phage-ssdAbs. A) Positive anti-TK1 phage-ssdAbs tested in monoclonal phage ELISA. Their capacity to bind TK1 and the stability of each clone was confirmed. B) Representative image showing the color development generated by the positive clones and plate layout indicating the position of each clone that was tested.

After confirming their binding to TK1, the positive clones were tested with dose calibrations curves to see if they were able to bind TK1 proportionally to its concentration and to determine their sensitivity. The 14 clones showed all sigmoidal curves according to our non-linear 4-point logistic analysis. The goodness of fit test showed R squares ranging between 0.9964-0.9772. The curves behaved according to receptor-ligand interactions models, showing that the binding of the anti-TK1-sdAbs were proportional to the concentration of TK1 in each well (Fig. 4A). This could also be visually appreciated in the colorimetric reactions in the dilution curves for each clone (Fig. 4B). There was not significant background signal produced by the clones in the blanks.

**Fig.4.**
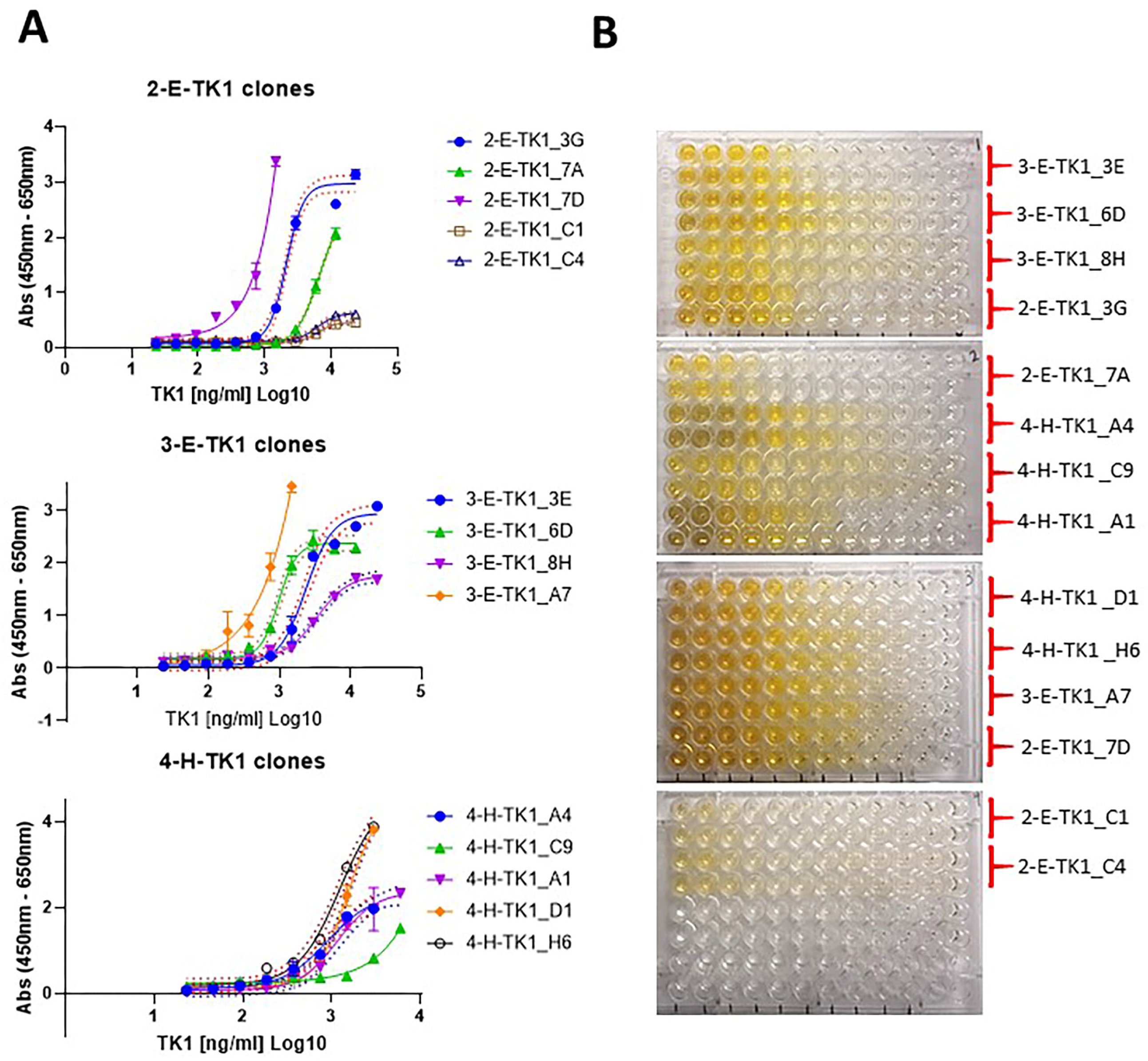
A) Dose response curves corresponding to anti-TK1 sdAb fragments obtained through various rounds of selection. The data was Log transformed and the curves were analyzed using a 4-parameter non-linear regression. B) A representative image of the colorimetric reaction showing that the signal of each anti-TK1 sdAb fragment is proportional to the concentration of TK1 protein. Negligible or no significant signal was produced in the blanks.

The clones that were the most sensitive were clones 4-H-TK1_A1 and 4-H-TK1_D1. Using the phage supernatant from these clones we were able to detect TK1 protein levels as low as 23 ng/ml in monoclonal phage ELISA (Fig. 5A). Further sequencing of the clones revealed that the sequences between clones 4-H-TK1_A1 and 4-H-TK1_D1 were different.

**Fig.5.**
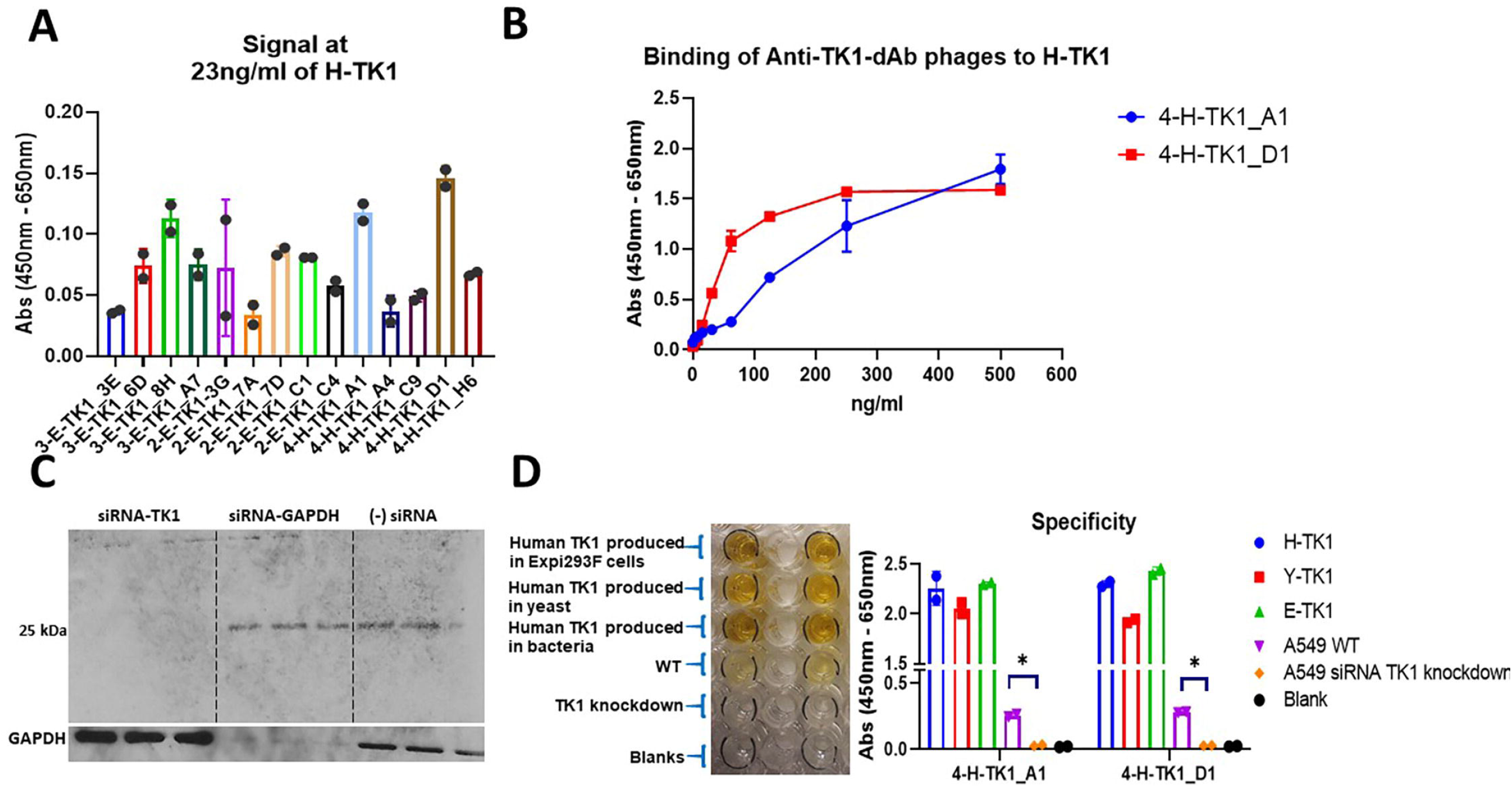
Sensitivity and specificity of the anti-TK1-sdAb fragments. A) The 14 anti-TK1 sdAb fragments were tested with a minimal fixed concentration of 23ng/ml of human TK1 produced in human cells (H-TK1). The sdAbs 4-H-TK1_A1 and 4-H-TK1_D1 produced the highest signals. B) Dilution curves with H-TK1 and the top two clones. Concentrations ranged from 500 ng/ml to 3.9 ng/ml, the fragments keep their binding properties after being expressed as sdAb fragments C) siRNA TK1 knock down validation. TK1 was knockdown in A549 cells. The knockdown was validated using commercial anti-TK1 antibody. D) Validation of the top 2 anti-TK1-ssdAbs. The binding capacity of the sdAb fragments was tested against 3 different sources of human recombinant TK1 and cell lysate from the cancer cell line A549 and cell lysate from A549 TK1 knockdown. It can be appreciated that both fragments bind to the 3 different recombinant TK1 proteins. A significant difference can be appreciated in the signal coming from the normal cell lysate in comparison with the TK1 knockdown cell lysate.

Anti-TK1-sdAbs phages 4-H-TK1_A1 and 4-H-TK1_D1 shown capacity to bind H-TK1 in phage ELISA. The signal was proportional to H-TK1 concentration (Fig. 5B). Validation of the clones 4-H-TK1_A1 and 4-H-TK1_D1 using a siRNA TK1 knockdown and different sources of recombinant human TK1 revealed that the clones were able to bind to E-TK1(produced in *E. coli*), H-TK1(produced in Expi293F cells) and TK1 produced in a yeast system. Moreover, the signal coming from cell lysate from A549 cells was significantly higher (∼10-fold) to the signal coming from the cell lysate of A549 TK1 knockdown for both 4-H-TK1_A1 (*P*<0.0296) and 4-H-TK1_D1 (*P*<0.0129). A TK1 knock down was produced as previously described for this experiment (Fig 5C and D) [27].

### Amplification of sdAB fragments, sequencing and cloning into pET-scFv-T vector

The dAb fragments were amplified by PCR and ligated into the pEt-scFv-T vector successfully as shown in Figure 6. The anti-TK1-sdAb vectors were sequenced, and the amino acid sequences were deduced from their nucleotide sequences (Table 2).

**Fig.6.**
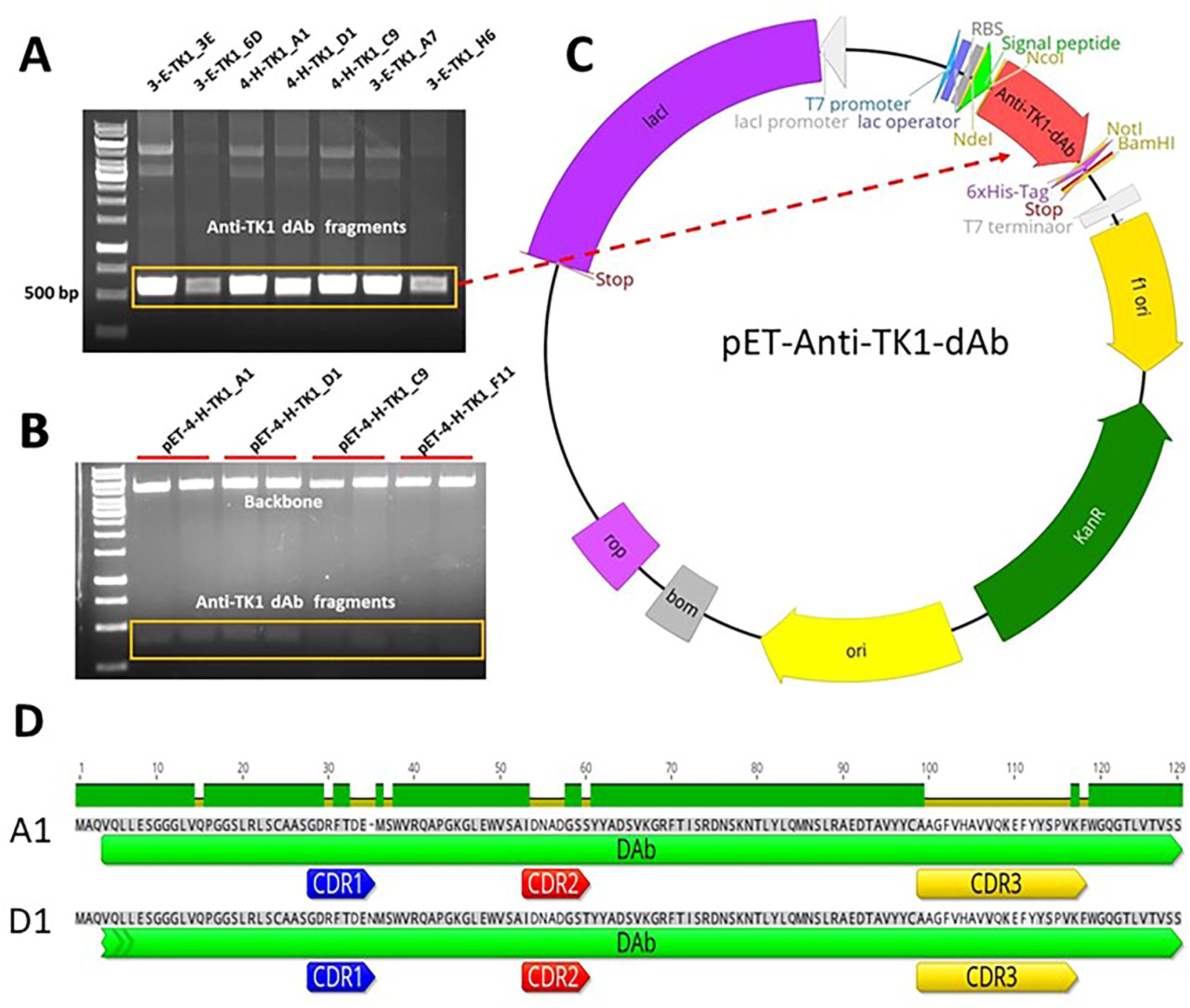
Amplification and ligation of anti-TK1-dAb fragments into the pET-scFv-T expression vector (Addgene #67843). A) a representative image of a PCR showing amplification of anti-TK1 dAb fragments. B) Restriction enzyme analysis with NcoI and NotI enzymes of pET-Anti-TK1-dAb constructs. C) Map of a pET-Anti-TK1-dAb construct. Fragments are ligated into the pET-scFv plasmid using NcoI and NotI restriction sites. D) Alignment of the 4-H-TK1_A1 and 4-H-TK1_D1 sdAb sequences shows the differences of the sdAbs are in their CDRs.

**Table 2.**
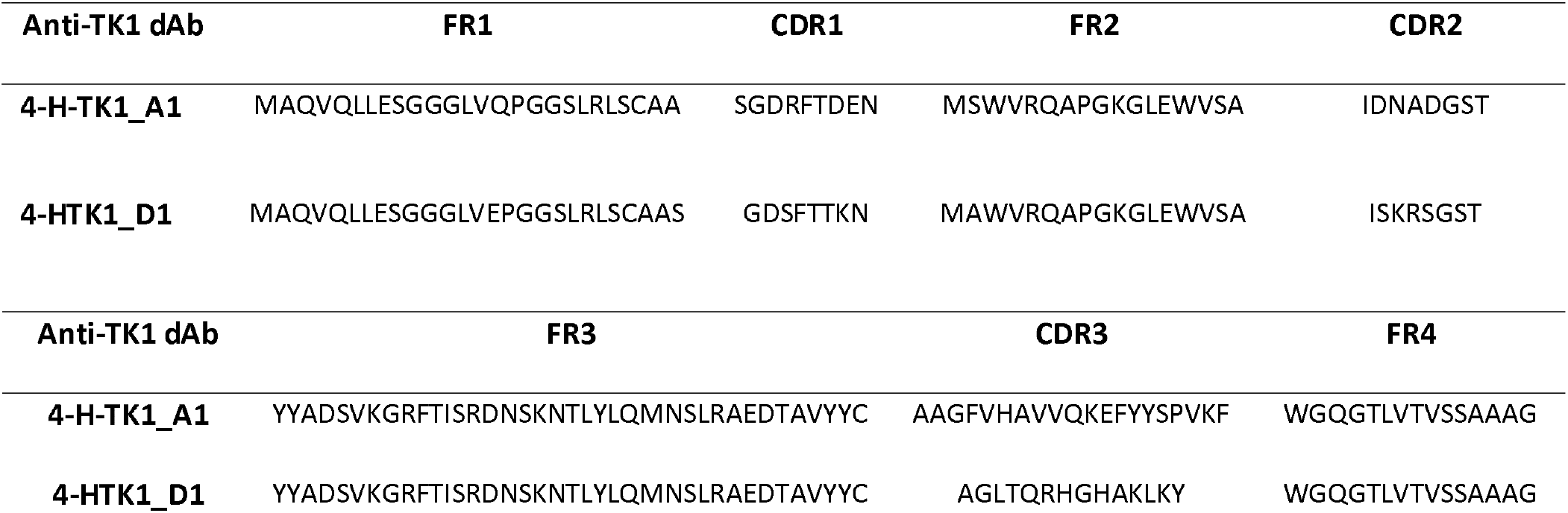
The deduced amino acid sequences of the top two anti-TK1 sdAb fragments isolated through phage display. CDRs were mapped using the IgBLAST tool. The antibodies sequences were aligned and compared using Genious prime software

Further analysis of the nucleotide sequences of the Anit-TK1 dAb fragments with the IgG blast tool from NCBI revealed the specific sites for their respective CDRs. The annotated sequences are found in Table 2. In addition, the alignment of the sequences using genious software confirmed that the annotated CDRs were the regions with most differences among the sequences while the heavy chain framework regions remained conserved (See Fig. 6D). This is consistent with the original description of the library which was built in a human VH framework and introduced diversity in the CDRs.

### Analysis of protein structure of Anti-TK1-sdAb fragments and modeling of sdAb-TK1 complexes

After submitting the corresponding amino acid sequences from the Anti-TK1 sdAb fragments A1, and D1 into the GalaxyTBM server, 5 model structures were generated for each fragment. The most stable strcuture from each anti-TK1-sdAb was then visualized using the VMD software. The Anit-TK1-sdAb fragments presented a typical structure of a sandwich of two antiparalel β-sheets according to previously reported single domain structures [40]. Their CDRs were contained in the loops conecting their antiparalel β-sheets having the CDR3 a longer loop according to the sdAb library design (Fig. 7 A-B). Furthermore, analysis of the interaction between the anti-TK1_4-H-A1 sdAb and the 1XBT crystal structure of TK1 [41] using the high ambiguity driven protein-protein docking (HADDOCK) web server revealed the interaction of the anti-TK1 sdAb 4-H-TK1_A1 through its CDRs with TK1. The CDRs seemed to interact with the α1-ribbon towards the N-terminus and two regions close to the β-ribbons at the c-terminus of the TK1 molecule (Fig. 7C and D).

**Fig.7.**
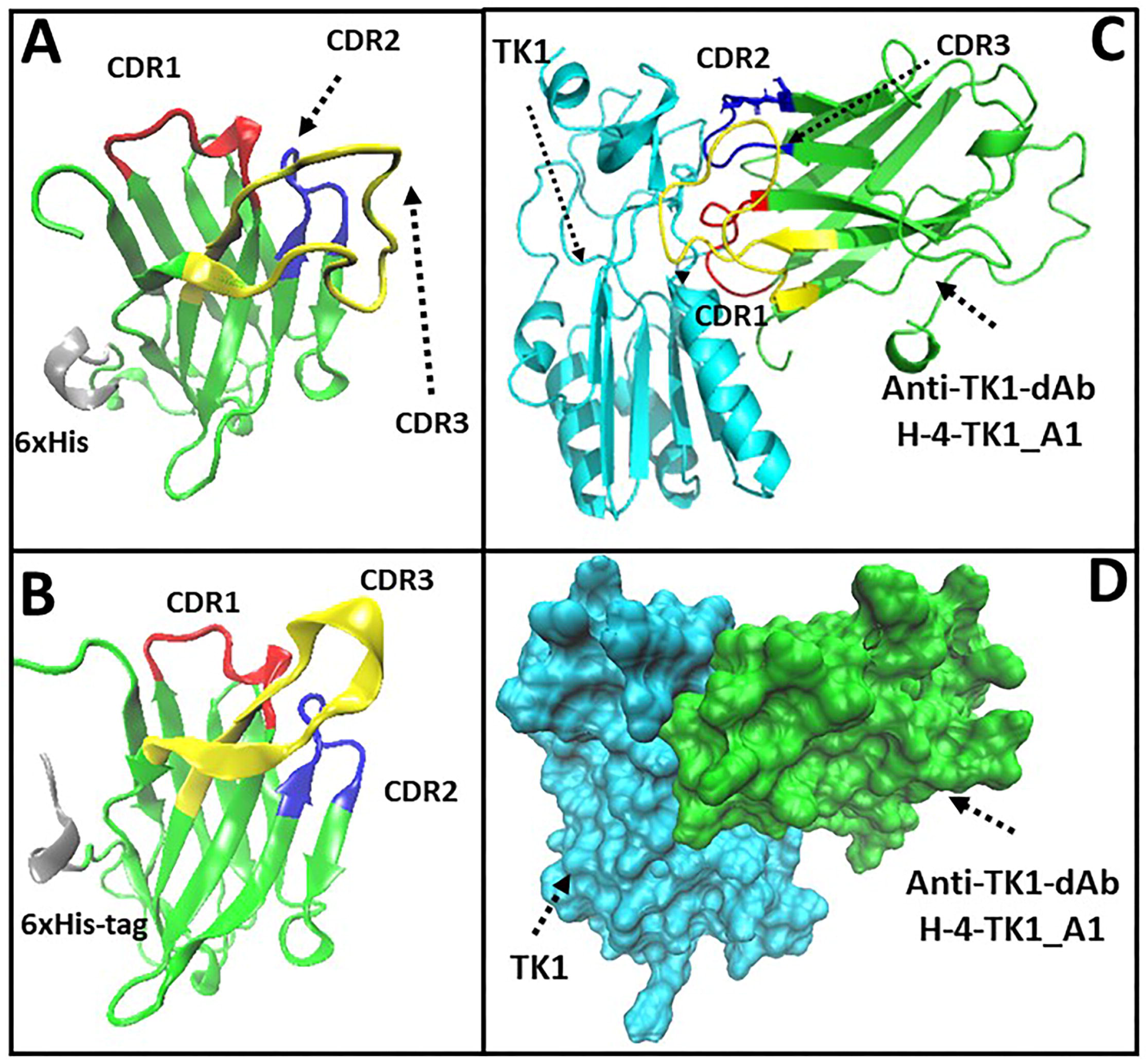
The 3D structure of the top two anti-TK1-sdAb fragments based on their deduced amino acid sequences. The Anti-TK1-sdAb fragments were modeled using GalaxyWeb TBM server. The most stable structures were then visualized using the VMD 1.9.3 software. CDRs were mapped by analyzing the anti-TK1 sdAb amino acid sequences with the IgBlast tool from NCBI. A) H-4-TK1_A1 sdAb. B) H-4-TK1_D1 sdAb. C) High ambiguity driven protein-protein docking analysis using the HADDOCK 2.4 webserver. The most stable structures of the H-4-TK1-A1 sdAb fragment and human TK1 protein monomer were analyzed to predict their protein-protein interactions. The analysis shows that the most reliable TK1-sdAb fragment complex would bind through its CDRs to TK1. The CDRs would interact with the α1-ribbon towards the N-terminus and two regions close to the β-ribbons towards the c-terminus of the TK1 molecule. D) Docking between the Anti-TK1 sdAb H-4-TK1_A1 and the monomer of human TK1 from the 1XBT crystal structure.

### Expression and characterization of purified sdAb fragments

The anti-TK1 sdAbs H-4-TK1_A1 and H-4-TK1_D1 were successfully expressed using the pET-scFv-T system. The fragments were detected with anti-His-HRP and anti-VSV-G-HRP antibodies in Western blot as could be observed (Fig. 8A). Moreover, coomassie blue staining showed successful purification of a 12-15 kDa band which matched the size of the band shown in Western blot. Purification of sdAb fragments with Ni-NTA bind-His resin yields were between 1 up to 4 mg/ml of purified fragment (Fig. 8B). Alternatively, the fragments that are VSV-G-tagged were purified using protein A columns (Fig. 8C). Although, the yields in protein A purification were lower, about 0.4 mg/ml (Fig. 8D).

**Fig.8.**
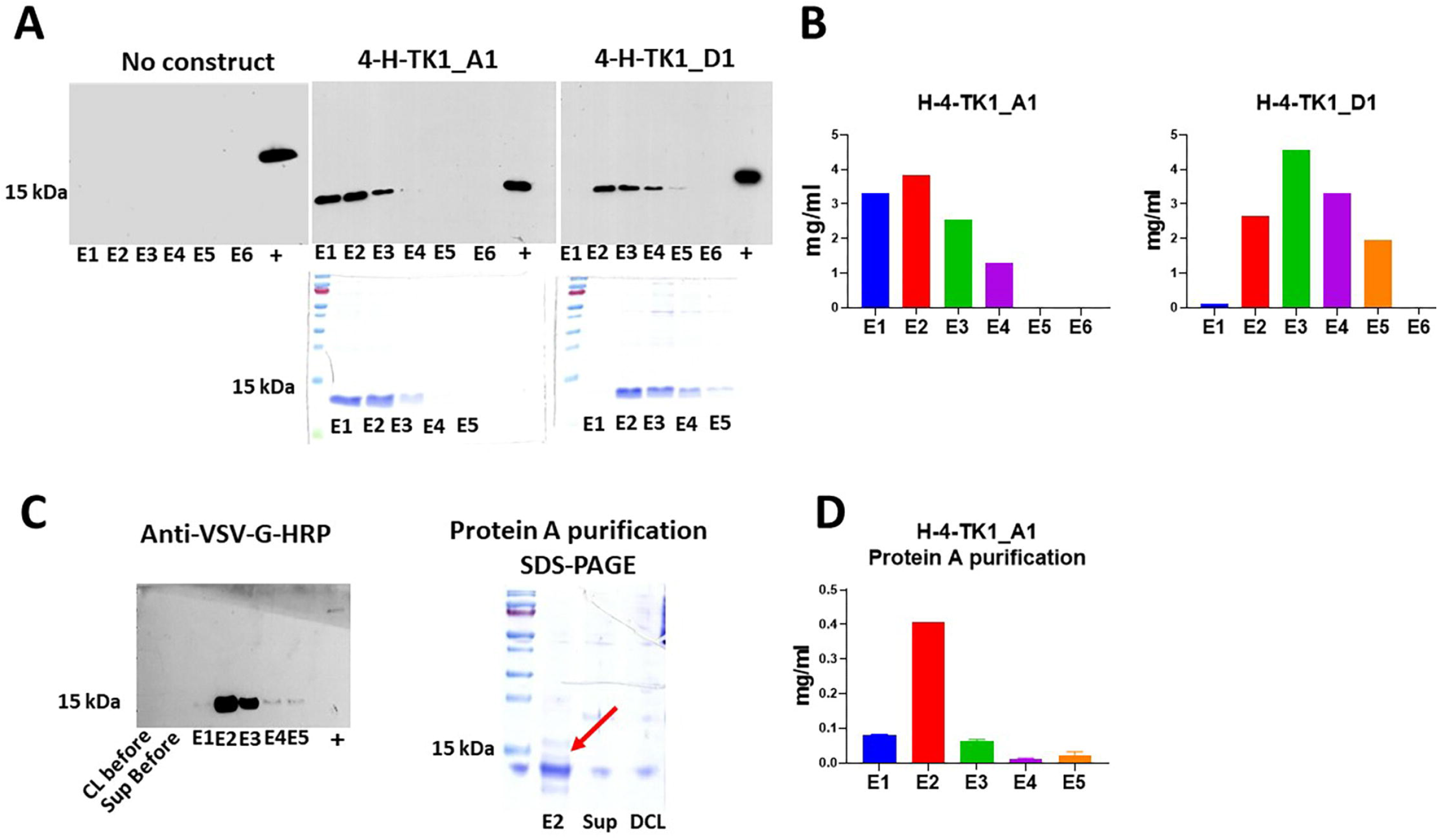
Expression and purification of anti-TK1 sdAbs. A) Detection of anti-TK1 sdAb fragments with anti-His-HRP antibody in Western blot and their respective SDS-PAGE analysis. B) Quantification of the His-tag purified sdAb fragments with BCA assay. The protein yields ranged between 1-4 mg/ml of purified sdAb. C) Western blot and SDS-PAGE analysis of protein A purified anti-TK1 sdAb H-4-TK1_A1. The antibody fragments can alternatively be purified with protein A purification. D) Quantification of protein A purified anti-TK1 sdAb H-4-TK1_A1.

### Western blot and soluble ELISA

After being expressed as sdAb fragments without a PIII gene fusion, the purified anti-TK1-sdAbs kept their binding properties to TK1 similarly to the ones observed when expressed as PIII fusions displayed on filamentous phages. These were tested through soluble ELISA. As shown, the anti-TK1 sdAbs bound proportionally to the concentration of TK1 in the wells. Concentrations ranged between 500 ng/ml-1.9 ng/ml of H-TK1. The anti-TK1-ssdAbs were able to produce signal significantly higher than the blanks at concentrations 3.9 ng/ml for H-4-TK1-A1 and 1.9 ng/ml for H-4-TK1-D1(Fig. 9A)

**Fig.9.**
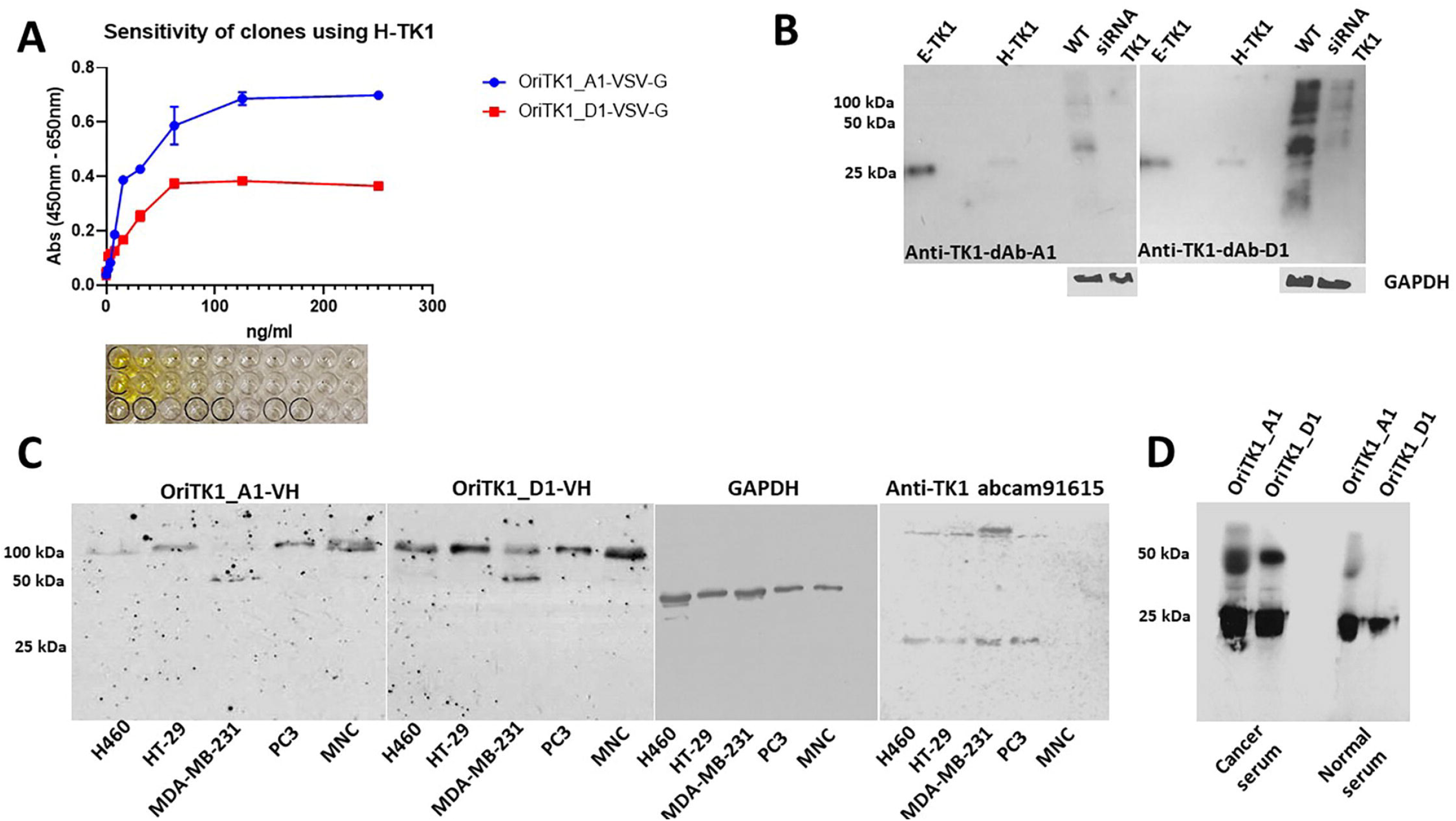
Soluble ELISA and Western blot analysis. A) Purified recombinant anti-TK1 sdAbs kept their binding properties H-TK1 after being expressed in *E. coli* and His-tag purified as observed in soluble ELISA. B) The anti-TK1 sdAbs showed binding to recombinant human TK1 produced in both *E*. coli (E-TK1) and human cells (H-TK1). The fragments were validated comparing their binding to TK1 in cell lysate of A549 cells (100 ug) and A549 TK1 knockdown (100ug). C)Detection of TK1 in cell lysate of 4 different cancer cell lines and normal MNCs (20ug each). The fragments were able to detect TK1 tetrameric form, controls included commercial anti-TK1 abcam91615 and GAPDH. D) A representative Western blot image showing detection of TK1 in human serum from a cancer patient and a healthy individual using anti-TK1_A1 and D1 fragments. For each cancer and healthy serum 10 ug of serum protein were loaded into each lane. A difference in the signal can visually be observed being the signal in the cancer serum slightly higher than the signal in normal serum

The anti-TK1-sdAbs were then tested in Western blot. As can be observed in Fig. 9B, the anti-TK1-ssdAbs showed binding to recombinant human TK1 produced in *E. coli*, and Expi293F cells. Moreover, the Anti-TK1 sdAb fragments showed a significantly higher signal in A549 cell lysate compared to A549 TK1 siRNA knockdown, particularly fragment 4-H-TK1_A1 showed specificity for TK1 (Fig. 9B).

Once we confirmed the binding of the fragments to purified recombinant TK1 and validated their specificity using a TK1 siRNA knockdown we proceeded to test its capacity to detect TK1 in cell lysates of different cancer cell lines, including, NCI-H460, HT-29, MDA-MB-231, PC3 and human MNC. It could be observed that the anti-TK1 sdAbs were able to detect bands corresponding 100 kDa in all cell lysates. In the case of MDA-MB-231 cells the sdAbs were also able to detect 50 kDa (Fig. 9C). This maybe corresponding to the active forms of TK1 dimer and tetramer. In a separate Western blot analysis, the anti-TK1-sdAbs were able to detect monomeric and dimeric forms of TK1 in human serum from a stage IV lung cancer patient which produce a higher signal compared to serum from a healthy patient (Fig. 9D).

### Flow cytometry

Detection of membrane expression of TK1 in NCI-H460 cells was done by staining the cells with PEG purified phages from each respective clone. We observed that after subtracting the background binding from the anti-M13-APC antibody there was an increase in binding to the cells stained with the anti-TK1-sdAbs of 20% and 25% for H-4-TK1_A1 and H-4-TK1_D1 sdAbs respectively (Fig. 10A). Therefore, the anti TK1 sdAbs showed capacity to detect membrane expression of TK1 on NCI-H460 cells. NCI-H460 cells were simultaneously screened with the commercial anti-TK1 antibody ab91651.

**Fig. 10.**
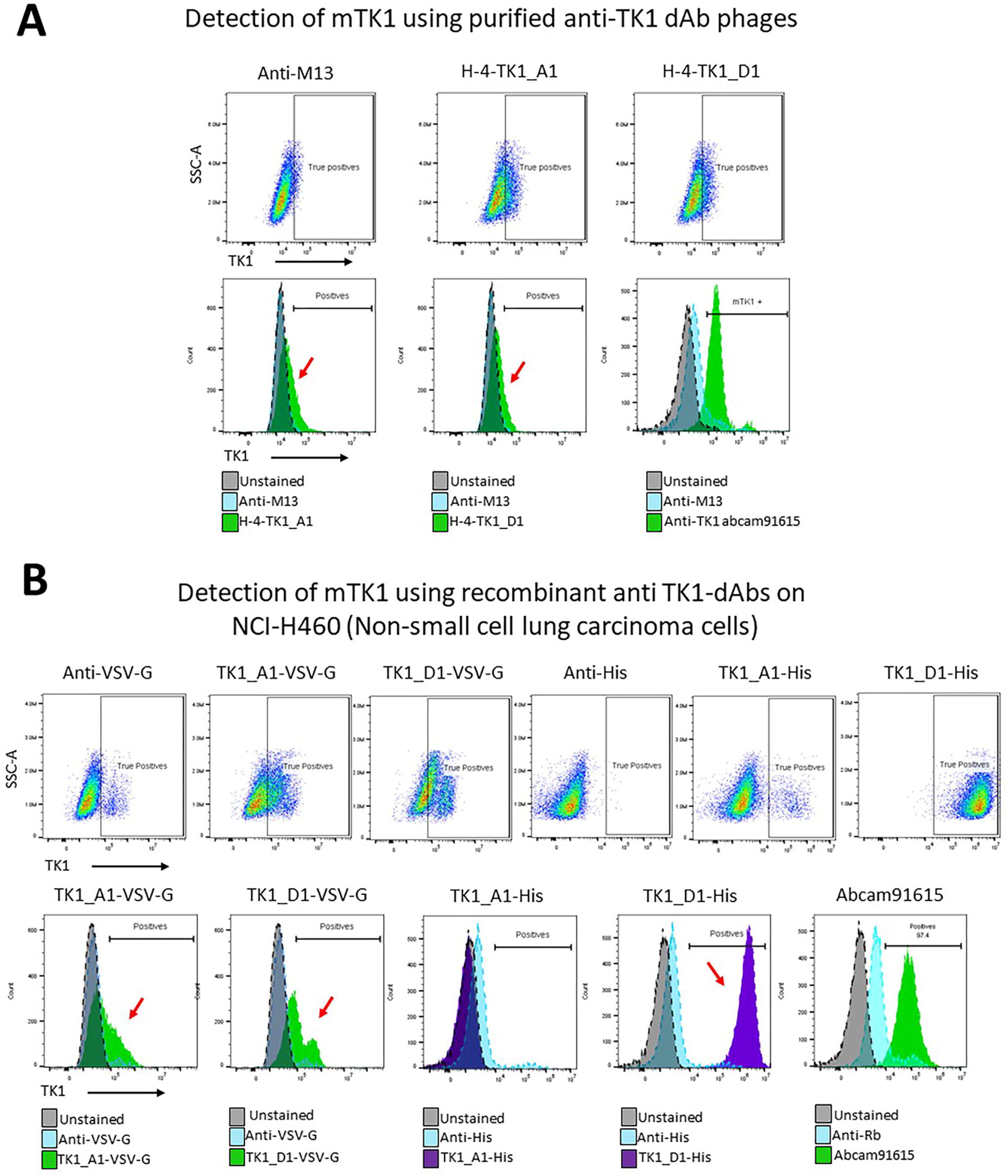
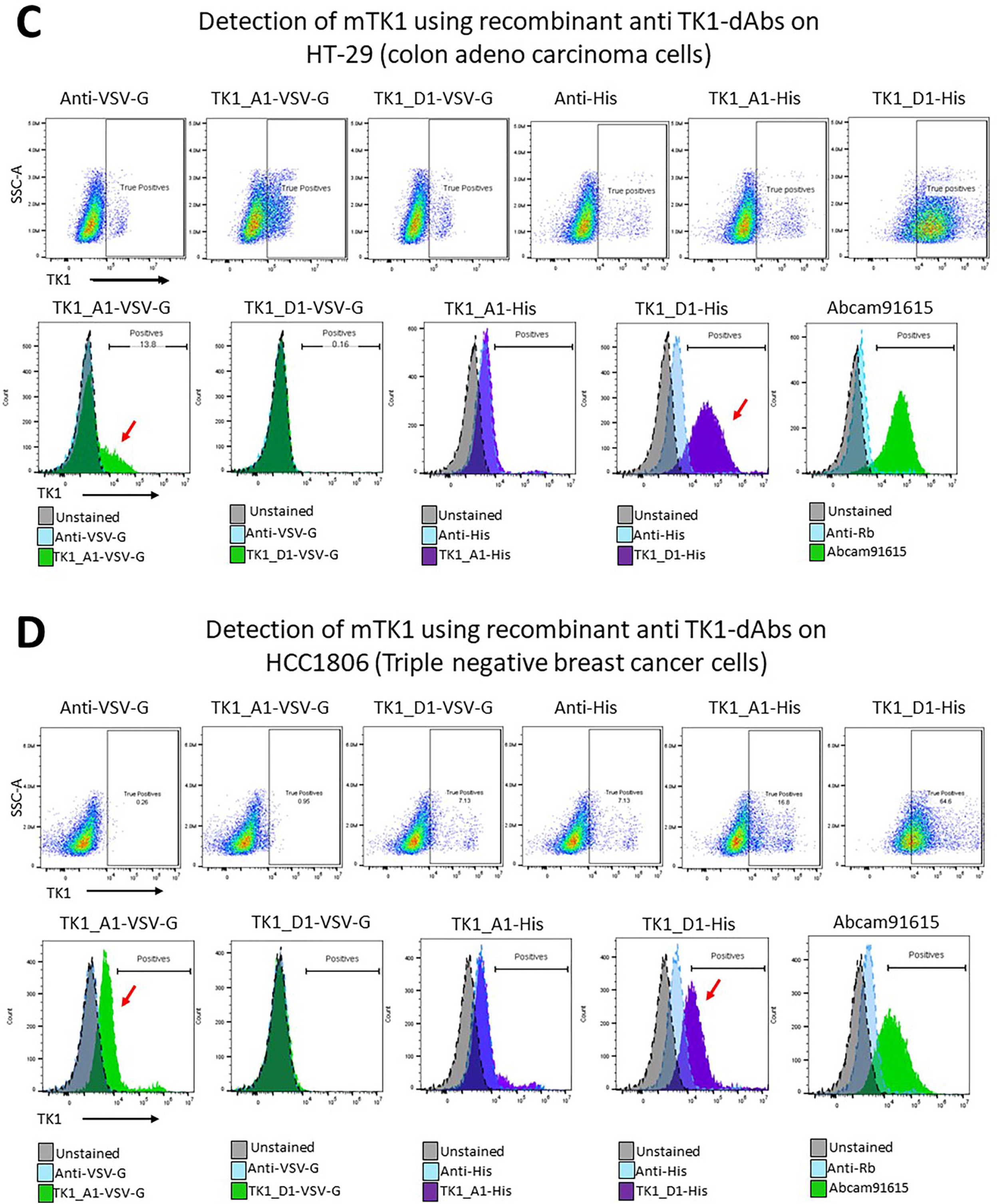
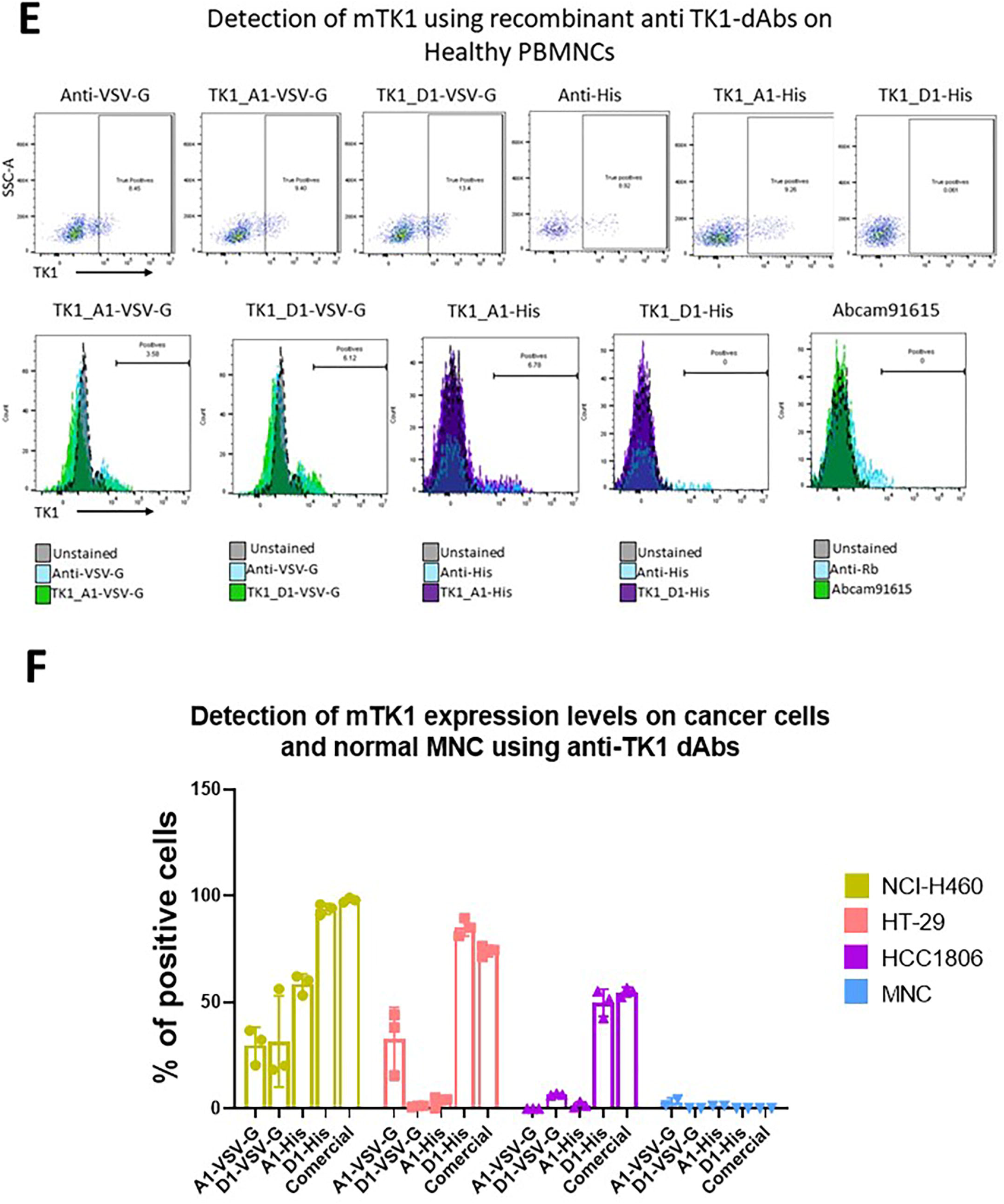
Detection of mTK1 through flow cytometry with anti-TK1 sdAb fragments on cancer cells and healthy MNC. A) NCI-H460 cells were stained with the purified phage-anti-TK1 sdAb fragments, a shift in the population could be observed using both phage-sdAb fragments. B-E) Expressed and purified anti-TK1-sdAb_A1 and D His-tagged and VSV-G tagged were used to stain NCI-H460 (lung), HT-29 (colon), HCC1806 (Triple negative breast cancer) and healthy lymphocytes. The level of mTK1 detection using the anti-TK1 sdAb fragments were comparable to the levels detected by commercial anti-TK1 antibody. The highest levels of mTK1 were detected on NCI-H460, following HT-29 and then HCC1806. No significant binding was detected on normal MNC with the anti-TK1 sdAb fragments nor with commercial anti-TK1 antibody. F) The different levels of mTK1 on cancer cells and variable binding of anti-TK1 sdAb fragments. Only A1-His sdAb showed consistent binding similar to commercial Ab.

After confirming the capacity of the TK1-sdAb-phages A1 and D1 to detect mTK1 on cancer cells the fragments were expressed as sdAbs and used to stain different cancer cell lines to test their ability to detect mTK1. The sdAbs were expressed as two different versions; His-tagged and VSV-G tagged. Among the 4 sdAbs, TK1-A1-His showed the most consistent binding to mTK1 on NCI-H460(∼95%), HT-29(∼87%) and HCC1806(∼53%) (Fig. 10 B-D, F). These expression levels were comparable to those seen with commercial TK1 antibody 97%, 72% and 55% for NCi-H460, HT-29 and HCC1806 respectively. Only in NCI-H460 cells which had the highest number of cells positive for mTK1 all the fragments His and VV-G tagged showed binding (Fig. 10B). D1-VSV-G fragment showed no binding in HT-29 cells and HCC1806 while A1-VSV-G and A1-His showed variable levels of binding in both cell lines. However, not as consistent as D1-His fragment. This may be due to differences in their tags, expression levels in each cell line and possible binding to different epitopes. No significant binding of the anti-TK1 sdAb fragments was found on normal lymphocytes after subtracting non-specific binding of secondary antibody anti-Human IgG FITC. No significant binding of the commercial anti-TK1 antibody was detected neither on normal lymphocytes (Fig 10E). It can be appreciated that the cancer cell lines expressed variable levels of mTK1 being NCI-H460 the one with the highest number of positive cells followed by HT-29 and HCC1806 (Fig. 10F).

### Cloning of the Anti-TK1-sdAb fragments into the pFUSE-IgG1e5 vector and expression of recombinant antibody in CHO.K1 cells

Before cloning the anti-TK1-sdAb sequences into an expression vector, site directed mutagenesis (SDM) was performed to change the amber stop codons present in the sdAb sequences to glutamic-acid. The anti-TK1-sdAb fragments were then successfully ligated into the pFUSE-IgGe5-IL2 expression vector at the NcoI and EcoRI restriction sites. Restriction analysis of these constructs showed the successful insertion of the sdAb genes into pFUSE-IgGe5-IL2 vector (Fig. 11A). After 72 and 96 hr of transfecting CHO.K1 cells with the pFUSE-4-H-TK1_A1 and D1, and a control without sdAb construct, the collected supernatant was run through a protein A column. The recombinant antibodies were successfully purified from supernatant (0.4-0.26 mg/ml). Western blot analysis and SDS page showed for both sdAb-Fc antibodies a ∼38-40 kDa band which is the expected size for the monomer of the anti-TK1-sdAb-IgG1 fusion. IgG1-no sdAb construct produced alone an engineered IgG1 construct of smaller size. The recombinant antibodies were all positive in Western blot to anti-human IgG antibody confirming the presence of the engineered IgG1 heavy chain fused to the sdAbs (Fig 11B). In addition, the recombinant antibodies showed capacity to detect mTK1 on cancer cells in flow cytometry (Fig 11C). A shift in the population of NCI-H460 cells could be appreciated when stained with the TK1-A1-IgG1 and D1 antibodies were used while no shift was detected using IgG1 fragment alone.

**Fig.11.**
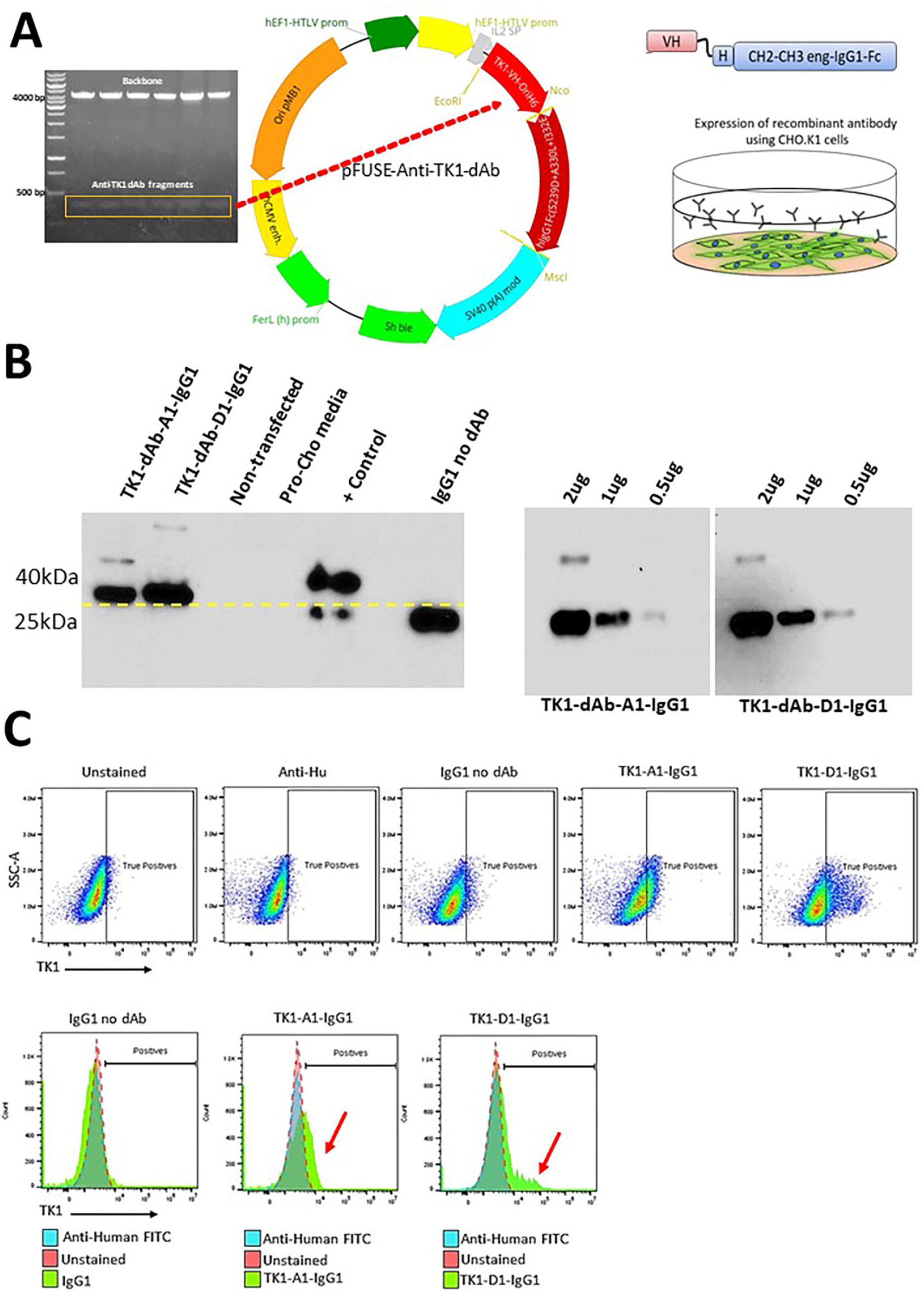
A) Construction of an Anti-TK1 sdAb fragment. Restriction analysis of pFUSE-anti-TK1-sdAb plasmids and their respective maps. B) Expression and detection of Anti-TK1-sdAb-IgG1 antibodies. As can be observed sdAb-IgG1 fusions produced higher molecular weight bands compared to IgG1-no sdAb constructs. No human IgG was detected in untransduced CHO.K1 cells. C) The anti-TK1-sdAb-IgG1 antibodies still detecting mTK1 in NCI-H460 cells. IgG1-no sdAb did not bind to NCI-H460 cells.

### Anti-TK1-sdAb antibodies elicited in vitro ADCC responses of human MNC against cancer cells expressing mTK1

To test the potential use of the Anti-TK1-sdAb antibodies for the immunotargeting of cancer cells that express mTK1. We co-cultured TK1 high-level expressing tumor cells with human MNC, added anti-TK1-sdAb-Fc antibodies and monitored the ADCC response through time. The NCI-H460 cell line was used as this cell lines previously showed to express the highest levels of mTK1 on the cell surface in flow cytometry (Fig. 11F). An initial test using different concentrations of Anti-TK1-ssdAbs showed that the cell killing was proportional to the concentration of antibody used (Fig. 12A) and it was found that a concentration of 10 μg/ml of the engineered anti-TK1 antibodies was necessary to cause a significant ADCC response. Cells treated with the anti-TK1-sdAb-IgG1_A1 and D1 antibodies at 10 μg/ml and co-cultured with human MNCs had a significant ADCC response against the cancer cells when compared to isotype (*P*<0.0395) and no antibody controls (*P*<0.0038) with the (Fig. 12B) anti-TK1-sdAb-IgG1_A1 after 88 hr. Although the 2-way ANOVA analysis did not show the difference was significant with the anti-TK1-sdAb-IgG1_D1 antibody a reduction of more than 50% of target cells was observed compared to Isotype controls. Imaging of the cells revealed that after 96 hr there was a difference in the cell health and number of MNC clustering targets cells. It can be visually appreciated that cells treated with anti-TK1-sdAb-IgG1 experienced a more severe ADCC response than thus treated with the isotype control (Fig. 12B). Statistical analysis at individual time points indicates that the percentage of cell killing is significantly higher at 96 hr when TK1 is targeted using the anti-TK1-sdAb-IgG1_A1 (*P*<0.0267) and D1 (*P*<0.0265) compared to controls and at 72 hr (*P*<0.0207 and *P*<0.0246 respectively) (Fig. 12C).

**Fig. 12.**
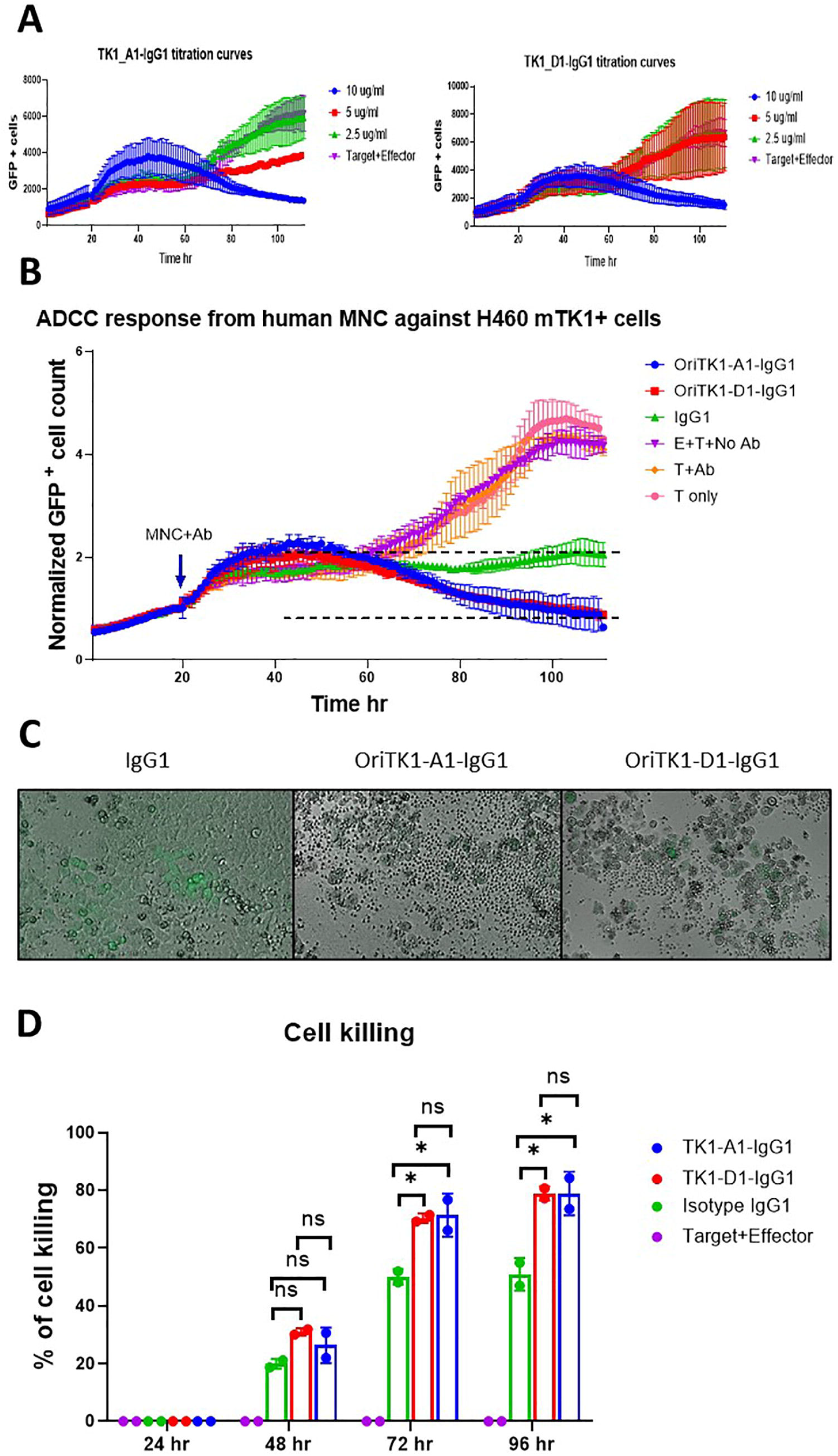
Anti TK1-sdAb-Fc antibodies elicit ADCC responses against NCI-H460 cells expressing mTK1. A) Optimization of several antibody concentrations. The decrease in GFP+ NCI-H460 cells co-cultured with MNC over time is proportional to the concentration of Anti-TK1-sdAb-IgG1/ml used. B) A significant ADCC response is elicited by MNCs against NCI-H460 cells when anti-TK1-sdAb-IgG1 antibodies are added compared to controls. After 96hr the cell killing can visually be appreciated to be more severe in the cells were treated with the anti-TK1-sdAb-IgG1 antibodies. C) Percentage of cell killing calculated every 24 hr. The percentage of cell killing is significantly higher after 72hr and 96hr when anti-TK1-ssdAbs are added in comparison to IgG1 isotype control and effector only-no sdAb control.

## Discussion

From its early beginnings’ studies involving TK1 have been focused mainly on its use as a tumor biomarker [42, 43]. However, new evidence has shown that TK1 may have an emerging role as a tumor target for cancer therapy. Recent studies suppressing the expression of TK1 in cancer cells have shown that the silencing of TK1 decreases the capacity of lung adenocarcinoma, pancreatic and thyroid carcinoma cells to proliferate, migrate or make mesenchymal transitions [22,23,44]. Additionally, some of the TK1 forms seems to be able to associate to the cell membrane of cancer cells, and event that apparently restricted to malignancy. Thus, it is clear that the development of therapeutics that can specifically target TK1 are necessary to explore the potential of TK1 as tumor target. Monoclonal antibodies are the fastest growing biopharmaceuticals in immuno-oncology [45]. Previously it has been shown that anti-TK1 monoclonal antibodies could be used for the immunotargeting of TK1 in several cancer types [27]. Although some monoclonal antibodies against TK1 have been developed there are currently no humanized versions of TK1 antibodies suitable for therapy [13, 27,46]. Moreover, the recent increase in the use of phage display antibody libraries have proven the advantages of using sdAb fragments in the development of therapeutic antibodies for cancer treatment. Single domain antibodies are characterized for their smaller size while keeping all the binding properties of full-length antibodies [47]. In this study anti-TK1 sdAbs were isolated from a sdAb library through phage display. This was the first time that the isolation and characterization of 100% human sdAb fragments specific for TK1 has been reported. This study also provides evidence that single domain antibodies or nanobodies can be used to target mTK1 on cancer cells, an antibody approach that has not been previously used for the targeting of TK1. Furthermore, this study shows evidence that TK1 sdAbs can be incorporated in engineered IgG1 antibody constructs to generate molecules for potential immunooncology applications.

It is important to mention that the process through which we selected the antibodies using TK1 produced in bacteria and TK1 produced in human cells was to make sure we obtain a suficient number of sdAbs and that the anti-TK1 sdAbs were able to bind properly folded human TK1. ELISA data in this study showed that the anti-TK1 sdAbs fit the receptor-ligand model and that the binding of the anti-TK1-sdAb fragments was dependent of the amount of available antigen. This study also shows that the anti-TK1-ssdAbs were able to be expressed without the PIII fusion while keeping their binding properties previously showed in phage monoclonal ELISA. Validation with an siRNA TK1 knockdown indicated that the antibody fragments developed were specific for human TK1. Furthermore, the flow cytometry data showed that the nanobodies can potentially be used to target cancer cells expressing TK1, particularly mTK1. This early evidence indicates that anti-TK1-ssdAbs could be used for the development of experimental TK1-based therapeutics such as anti-TK1-sdAb fragments that could be conjugated with toxins or gold nanoparticles for anticancer photothermal therapy [48, 49]. Moreover, the anti-TK1-sdAb fragments could also be used for the development of immuno-oncology therapeutics such as engineered antibodies or chimeric antigen receptors [50].

As this study has shown, anti-TK1-sdAb-IgG1 antibodies are capable of targeting mTK1 on cancer cells and elicit an ADCC response of human MNC against TK1 high-level expressing cancer cells building up on previous findings. Contrary to previous anti-TK1 antibodies generated through conventional hybridoma technology, these engineered antibodies are completely human, are significantly smaller than full length antibodies and have an engineered IgG1 to enhance the ADCC response. Thus, they can have better tumor penetration than conventional antibodies, and can be used to better engage the immune system with tumor cells. Although it remains unclear why TK1 is localized to the cell surface of multiple cancer cell lines, the flow cytometry data and ADCC results described here suggest that it could be feasible to harness the immune system against tumor cells expressing mTK1. It is not the first time that a protein thought to be limited to the interior of the cell has been reported to be on the cell surface. Examples can be found in the HSP70 family of proteins. Although HSP70 proteins were thought to be limited to function only inside cells, it is well documented that they are secreted and localized on the cell membrane of cancer cells [51-52] Recent studies have shown that TK1 is present in exosomes [26]. Like HSP70 proteins a possible explanation could be that it is transitorily localized on the cell membrane as exosomes exit the cell fusing to the cell membrane, and through non-conventional protein-protein interactions. Moreover, it is also important to point out that other nucleotide salvage pathway enzymes have been reported to be localized on the cell membrane such as hypoxanthine-phosphoribosyl transferase (HPRT). Targeting of this enzyme with monoclonal antibodies has also been recently reported [53].

## Conclusion

This study reports the isolation and evaluation of human single domain antibodies against TK1 for the targeting of the tumor proliferation biomarker TK1 in lung, colon and breast cancer cells. The antibody fragments have potential as diagnostic and therapeutic agents. Although additional *in vivo* studies are required to confirm their efficacy. The antibody fragments can be successfully incorporated into IgG1 Fc constructs for the production of completely human engineered antibodies able to elicit significant ADCC responses from human MNC against cancer cells expressing mTK1.

The antibody fragments potentially can be used in other therapies such as chimeric antigen receptors (CAR) for T cells or other recombinant antibody constructs. The use of TK1 as a tumor target will enable the testing of experimental TK1-based therapeutics.

## Data Availability

All the relevant information is contained within this manuscript. Additional files regarding this study will be available from the authors upon a reasonable request basis

## Declarations

### Ethics approval and consent to participate

Isolation of human MNCs and serum from blood was obtained under approval of the Brigham Young University Institutional Review Board number 1734.

### Consent for publication

Personal information details from individual that donated blood were not included in this manuscript

### Authors’ contributions

EJV and KLO conceived the original scientific questions and experimental design of this research. EJV was the major contributor in the collection and analysis of the data of this manuscript. JDC, TBH, TOM, DMB, JRS and KRS assisted in the selection and evaluation of single domain antibodies, Western blot, ELISA, purification of antibodies and cell culture. EJV, KLO, RAR and SKW Contributed to the interpretation of the data and writing of this manuscript.

## Acknowledgments

We acknowledge the valuable contributions of Naomi Raper, Kelsey Bhingham, and Zachary Ewel for their technical support during the early stages of this research.

## References

1. Buj R and Aird MK. Deoxyribonucleotide Triphosphate Metabolism in Cancer and Metabolic Disease. Front endocrinol. 2018;9:177. doi: 10.3389/fendo.2018.00177

2. Traut TW. Physiological concentrations of purines and pyrimidines. Mol cell biochem. 1994;140(1);1-22. https://doi.org/10.1007/BF00928361

3. Villa E, Ali SE, Sahu U, Sahra IB. Cancer cells tune the signaling pathways to empower de novo synthesis of nucleotides. Cancers. 2019;11(5):688. doi: 10.3390/cancers11050688

4. Loffler M, Fairbanks LD,Zameitat E, Marinaki AM, Simmonds HA. Pyrimidine pathways in health and disease. TRENDS mol med. 2005;11:9. doi:10.1016/j.molmed.2005.07.003.

5. Somyajit K, Gupta R, Sedlackova H, Neelsen KJ, Ochs F, Rask MB, Choudhary C, Lukas J. Redox-sensitive alteration of replisome architecture safeguards genome integrity. Science. 2017;358:797–802

6. Munch-Petersen B, Cloos L, Jensen HK and Tyrsted G. Human Thymidine Kinase 1 regulation in normal and malignant cells. Advan. Enzyme Regul. 1995;35:68–69

7. Hanahan D, Weinberg RA. Hallmarks of cancer: the next generation. Cell. 2011;144:646–674. doi: 10.1016/j.cell.2011.02.013.

8. Birringe MS, Perozzo R, Kut E, Stillhart C, Surber W, Scapozza L, Folkers G. High-level expression and purification of human thymidine kinase 1 : Quaternary structure, stability, and kinetics. Protein expression purify. 2006;47:506–515. doi:10.1016/j.pep.2006.01.001

9. Zhou J, He E, Skog S. The proliferation marker thymidine kinase 1 in clinical use. Mol Clin Oncol. 2013;1:18–28. doi: 10.3892/mco.2012.19

10. Ke, P. Y., & Chang, Z. F. (2004). Mitotic degradation of human thymidine kinase 1 is dependent on the anaphase-promoting complex/cyclosome-CDH1-mediated pathway. Molecular and cellular biology, 24(2), 514–526. https://doi.org/10.1128/mcb.24.2.514-526.2004

11. Kauffman MG, Kelly TJ. Cell cycle regulation of thymidine kinase: residues near the carboxyl terminus are essential for the specific degradation of the enzyme at mitosis. Mol Cell Biol. 1991 11:2538–2546. doi: 10.1128/mcb.11.5.2538

12. Gatt ME, Goldschimdt N, Kalichman I, Friedman M, Arronson AC, Barak V. Thymidine Kinase Levels Correlate with Prognosis in Aggressive Lymphoma and Can Discriminate Patients with a Clinical Suspicion of Indolent to Aggressive Transformation. Anticancer Res. 2015;35:3019–3026.

13. Jagarlamudi KK, Aronsson AC, Pilko G, Zupan M, Kumer K, Fabjan T. A clinical evaluation of the TK 210 ELISA in sera from breast cancer patients demonstrates high sensitivity and specificity in all stages of disease. Tumor Biology. 2016;37:11937–11945. doi: 10.1007/s13277-016-5024-z

14. Wang Y, Jiang X, Wang S, Yu H, Zhang T, Xu S, Li W, He E and Skog S. Serological TK1 predict pre-cancer in a routine health screening of 56, 178 people. Cancer Biomark. 2018;22(2):237–247. DOI: 10.3233/cbm-170846

15. Alegre MM, Weyant MJ, Bennett DT, Yu JA, Ramsden MK, Elnaggar A, Robison RA, O’Neill KL. Serum detection of thymidine kinase 1 as a means of early detection of lung cancer. Anticancer Res. 2014;34(5), 2145–51.

16. Alegre, MM, Robison, RA, O’Neill K L. Thymidine Kinase 1 Upregulation is an Early Event in Breast Tumor Formation. J. of Oncol. 2012;2012:1–5.

17. Chen ZH, Huang SQ, Wang Y, Yang AZ, Wen J, Xu XH, Chen Y, Chen QB, Wang YH, He E, Zhou J, Skog S. Serological thymidine kinase 1 is a biomarker for early detection of tumours-a health screening study on 35,365 people, using a sensitive chemiluminescent dot blot assay. Sensors. 2011;11(12):11064–80. doi: 10.3390/s111211064

18. Chen Y, Ying M, Chen YS, Hu M, Lin Y, Chen D, Li X, Zhang M, Yun X, Zhou J, He E, Skog S. Serum thymidine kinase 1 correlates to clinical stages and clinical reactions and monitors the outcome of therapy of 1,247 cancer patients in routine clinical settings. Int. J. of Clin. Oncol. 2010;15(4), 359–368.

19. Hallek M, Wanders L, Strohmeyer S, Emmerich B. Thymidine Kinase 1: a tumor marker with prognostic value for non-Hodgkin’s lymphoma and a broad range of potential clinical applications. Ann. of hematol. 1992;65(1):1–5.

20. He Q, Fornander T, Johansson H, Johansson U, Hu GZ, Rutqvist LE, Skog S. Thymidine Kinase1 in serum predicts increased risk of distant or loco-regional recurrence following surgery in patients with early breast cancer. Anticancer Res. 2006;26:4753–4760.

21. Jagarlamud KK, Shaw M. Thymidine kinase 1 as a tumor biomarker: Technical advances offer new potential to an old biomarker. Biomark. in Med. 2018;12:1035–1048. doi: 10.2217/bmm-2018-0157

22. Malvi P, Janostiak R, Nagarajan A, Cai G, Wajapeyee N (2019) Loss of thymidine kinase 1 inhibits lung cancer growth and metastatic attributes by reducing GDF15 expression. PLoS Genet 15(10): e1008439. https://doi.org/10.1371/ journal.pgen.1008439

23. Zhu X, Shi C, Peng Y, Yin L, Tu M, Chen Q, Hou C, Li Q and Miao Y. Thymidine Kinase 1 silencing retards proliferative activity of pancreatic cancer cell via E2F1-TK1-P21 axis. Cell proliferat. 2018;51:e12428.

24. Weagel EG, Meng W, Townsend MH, Velazquez EJ, Brog RA, Boyer MW, Weber SK, Robison RA, O’Neill KL. Biomarker analysis and clinical relevance of TK1 on the cell membrane of Burkitt’s lymphoma and acute lymphoblastic leukemia. OncoTargets and Therapy. 2017;10:4355–4367.

25. Weagel EG, Burrup W, Kovtun R, Velazquez EJ, Felsted AM., Townsend MH, Zachary EE., Suh E, Piccolo SR, Weber SK, Robison RA, O’Neill KL. Membrane expression of thymidine kinase 1 and potential clinical relevance in lung, breast, and colorectal malignancies. Cancer Cell Int. 2018;18:135.

26. Shojai TM. The mechanism of TK1 secretion in cancer cells (unpublished thesis). Swedish University of Agricultural Sciences, Upssala, Sweden.

27. Velazquez EJ, Brindley TD, Shrestha G, Bitter EE, Cress JD, Townsend MH, Berges BK, Robison RA, Weber KS, O’Neill KL. Novel monoclonal antibodies against thymidine kinase 1 and their potential use for the immunotargeting of lung, breast and colon cancer cells. 2020;20:127. https://doi.org/10.1186/s12935-020-01198-8

28. Parakh S., King D., Gan H.K., Scott A.M. (2020) Current Development of Monoclonal Antibodies in Cancer Therapy. In: Theobald M. (eds) Current Immunotherapeutic Strategies in Cancer. Recent Results in Cancer Research, vol 214. Springer, Cham. https://doi.org/10.1007/978-3-030-23765-3_1

29. Lu, R., Hwang, Y., Liu, I. et al. Development of therapeutic antibodies for the treatment of diseases. J Biomed Sci 27, 1 (2020). https://doi.org/10.1186/s12929-019-0592-

30. Manis JP. Overview of therapeutic antibodies. 2020. Accessed [2020 May 3] https://www.uptodate.com/contents/overview-of-therapeutic-monoclonal-antibodies#H26

31. Almagro JC, Pedraza-Escalona M, Arrieta HI, Perez-Tapia SM. Phage display libraries for antibody therapeutic discovery and development. Antibodies. 2019;8:44. doi:10.3390/antib8030044

32. Zhao A, Tohidkia MR, Siegel DL, Coukos G, Omidi Y. Phage antibody display libraries: a powerful antibody discovery platform for immunotherapy. Crit. Rev. Biotechnol. 2016;36(2):276–289. doi: 10.3109/07388551.2014.958978

33. Barderas, R., Benito-Peña, E. The 2018 Nobel Prize in Chemistry: phage display of peptides and antibodies. Anal Bioanal Chem 411, 2475–2479 (2019). https://doi.org/10.1007/s00216-019-01714-4

34. Frenzel A, Schirmann T, Hust M. Phage display-derived human antibodies in clinical development and therapy. MABS. 2016;8(7):1177–1194. http://dx.doi.org/10.1080/19420862.2016.1212149

35. Marintcheva B. (2018) Phage display. Harnessing the power of viruses. (pp 133–160). https://doi.org/10.1016/B978-0-12-810514-6.00005-2

36. Lee, C. M., Iorno, N., Sierro, F., & Christ, D. (2007). Selection of human antibody fragments by phage display. Nature protocols, 2(11), 3001.

37. Genious Prime 2019.0.3. https://www.geneious.com.

38. Humphrey, W., Dalke, A. and Schulten, K., “VMD - Visual Molecular Dynamics”, J. Molec. Graphics, 1996, vol. 14, pp. 33–38.

39. G.C.P van Zundert, J.P.G.L.M. Rodrigues, M. Trellet, C. Schmitz, P.L. Kastritis, E. Karaca, A.S.J. Melquiond, M. van Dijk, S.J. de Vries and A.M.J.J. Bonvin (2016). “The HADDOCK2. 2 webserver: User-friendly integrative modeling of biomolecular complexes.” J. Mol. Biol., 428, 720–725 (2015).

40. Alberts B, Johnson A, Lewis J, et al. Molecular Biology of the Cell. 4th edition. New York: Garland Science; 2002. The Shape and Structure of Proteins. Available from: https://www.ncbi.nlm.nih.gov/books/NBK26830/

41. Welin M, Kosinska U, Mikkelsen N, Carnrot C, Zhu C, Wang L, Eriksson S, Munch-Petersen B, and Eklund H. Structures of thymidine kinase 1 of human and mycoplasmic origin. PNAS. 2004;101:17970–17975

42. ONeill KL, Buckwalter M, Murray BK. “Thymidine kinase: diagnostic and prognostic potential”. Expert Rev. Mol. Diagn. 1 (4): 428–33. doi:10.1586/14737159.1.4.428

43. Ondrej Topolcan & Lubos Holubec Jr. The role of thymidine kinase in cancer diseases, Expert Opinion on Medical Diagnostics, 2:2, 129–141, DOI: 10.1517/17530059.2.2.129

44. Liu C, Wang J, Zhao L, He H, Zhao P, Peng Z, Liu F, Chen J, Wu W, Wang G and Dong F. Knockdown of Thymidine Kinase 1 Suppresses Cell Proliferation, Invasion, Migration, and Epithelial–Mesenchymal Transition in Thyroid Carcinoma Cells. Front. Oncol. 9:1475. doi: 10.3389/fonc.2019.01475

45. To cite this article: Philippe Valadon, Sonia M. Pérez-Tapia Renae S. Nelson, Omar U. Guzmán- Bringas, Hugo I. Arrieta-Oliva, Keyla M. Gómez-Castellano, Mary Ann Pohl & Juan C. Almagro (2019) ALTHEA Gold Libraries™: antibody libraries for therapeutic antibody discovery, mAbs, 11:3, 516–531, DOI: 10.1080/19420862.2019.1571879

46. US patent. US 2010/0173329 A1

47. Schrankel CS, Gökirmak T, Lee CW, Chang G, Hamdoun A. Methods in cell biology. Chapter 14-generation, expression and utilization of single-domain antibodies for in vivo protein localization and manipulation in sea urchin embryos. Elsevier 2019;151:353–376.

48. Van de Broek B, Devoogdt N, D’Hollander A, et al. Specific cell targeting with nanobody conjugated branched gold nanoparticles for photothermal therapy. ACS Nano. 2011;5(6):4319–4328. doi:10.1021/nn1023363

49. Yu, Y., Li, J., Zhu, X., Tang, X., Bao, Y., Sun, X., Huang, Y., Tian, F., Liu, X., & Yang, L. (2017). Humanized CD7 nanobody-based immunotoxins exhibit promising anti-T-cell acute lymphoblastic leukemia potential. International journal of nanomedicine, 12, 1969–1983. https://doi.org/10.2147/IJN.S127575

50. Li N, Fu H, Hewitt SM, Dimitrov DS, Ho M (August 2017). “Therapeutically targeting glypican-2 via single-domain antibody-based chimeric antigen receptors and immunotoxins in neuroblastoma”. Proceedings of the National Academy of Sciences of the United States of America. 114 (32): E6623–E6631. doi:10.1073/pnas.1706055114

51. Vega, V. L., Rodríguez-Silva, M., Frey, T., Gehrmann, M., Diaz, J. C., Steinem, C., Multhoff, G., Arispe, N., & De Maio, A. (2008). Hsp70 translocates into the plasma membrane after stress and is released into the extracellular environment in a membrane-associated form that activates macrophages. Journal of immunology (Baltimore, Md. : 1950), 180(6), 4299–4307. https://doi.org/10.4049/jimmunol.180.6.4299

52. De Maio A. (2014). Extracellular Hsp70: export and function. Current protein & peptide science, 15(3), 225–231. https://doi.org/10.2174/1389203715666140331113057

53. Townsend, M. H., Bennion, K. B., Bitter, E. E., Felsted, A. M., Robison, R. A., & O’Neill, K. L. (2021). Overexpression and surface localization of HPRT in prostate cancer provides a potential target for cancer specific antibody mediated cellular cytotoxicity. Experimental cell research, 403(1), 112567. Advance online publication. https://doi.org/10.1016/j.yexcr.2021.112567

